# A Method for Monitoring the Effective Air Change Rate for Respiratory Aerosols Using Real-Time Tracers

**DOI:** 10.1101/2023.05.05.23289521

**Authors:** Saurabh Pathak, Kalyan Kottapalli, Joshua L. Santarpia, Aaron D. Botham, Sam D. Molyneux, Jamie Balarashti

**Author notes:** **FDA Compliance:**This study was conducted in compliance with FDA Good Laboratory Practices (GLP) as defined in 21 CFR, Part 58.

## Abstract

Ventilation is one of the most critical components in a layered approach toward reducing the spread of airborne infectious diseases in indoor spaces. However, building ventilation systems act together with natural ventilation, local filtration systems and other aerosol removal processes to remove infectious aerosols from an occupied space. Airflow-based determinations of ACH do not account for the full range of aerosol removal processes; however understanding the effective aerosol removal rate is critical to providing airborne infection control.

In this study, we investigated the relationship between the calculated air change rate of a space (i.e. volumetric airflow based) and the effective air change rate for aerosol particle removal within the breathing zone based on direct measurements of the rate of change in tracer particle concentrations at representative occupant locations in a room. Further, we examined positional effects under well mixed and non-well mixed conditions.

Our results demonstrate that tracer particles combined with real-time sensors can be used to make rapid, accurate measurements of the effective air change rate (eACH) for respiratory aerosols within the breathing zone of non-well mixed rooms. We used two experimental test beds for these analyses. First, numerical simulation (computational fluid dynamic simulation, CFD) was conducted to visualize airflow and particle removal paths within a realistic large room. Here, simulated sensors were placed in concentric zones around a nebulizer providing test-particle releases. This CFD model allowed a direct comparison of the differences between eACH and airflow ACH values under varying levels of mixing and airflow, in a fully controlled system.

We then recapitulated this system in physical space to validate the CFD results under real-world conditions that include all mechanisms of particle removal that contribute to true aerosol clearance rates, including deposition and leakage. Here, we measured eACH using the decay of DNA tracer aerosols nebulized and monitored in real-time. We find that a standard sampling time of 15 minutes from the end of nebulization is sufficient to produce an accurate eACH value under non-well mixed conditions. The availability of a rapid direct test for eACH will enable empirical optimization of a wide range of ventilation and filtration mechanisms to reach and maintain target aerosol clearance rates that deliver reliable airborne infection control in typical indoor environments.

## 1. Introduction

The global health pandemic of 2020, driven by SARS-CoV-2, the virus that causes COVID-19, has challenged individuals and organizations to re-think everyday tasks. As the world settles into the post-pandemic life, businesses and organizations are re-evaluating their protections for minimizing the indoor spread of respiratory infections of all types among customers and employees. Critical to this is understanding how SARS-CoV-2 and other respiratory viruses are transmitted through the air within a facility. The transmission of SARS-CoV-2 occurs primarily through aerosols generated during breathing, speaking, coughing, or sneezing, which may land in the nose, mouth, or eyes [1]. In this context, aerosol particles can be considered the continuum of particles ranging from small droplet nuclei to large droplets. Various proportions of large and small aerosol particles are emitted from the individual depending on the type of activity performed, such as talking, coughing, or sneezing [2]. Although large respiratory droplets tend to fall out of the air quickly, aerosols containing smaller particles can travel across greater distances [3, 4].

Once infectious particles are exhaled, they move outward from the source. Two principal processes determine the amount of virus a person is exposed to following exhaled viral emission from an infected individual. First, the decreasing concentration of virus in the air as larger and heavier respiratory droplets containing virus fall to surfaces under the force of gravity, while very fine droplets and particles that remain in the airstream progressively mix with and become diluted within the air space. Second, the progressive loss of viral viability and infectiousness over time as the particles are influenced by environmental factors such as temperature, humidity, and ultraviolet radiation (e.g., sunlight) [6]. Ensuring proper replacement with clean, non-infectious air (outdoor, filtered or treated) reduces the concentration of airborne pathogens, providing effective infection control. Proper ventilation also reduces surface contamination by removing a portion of virus particles before they fall out of the air and settle on surfaces. In general, the greater the number of people in an indoor environment, the greater the need for ventilation.

ASHRAE is an international organization that establishes current ventilation standards for most indoor spaces. These standards have been designed to dilute bio-effluents (i.e., odors from people) and achieve basic levels of CO2 removal rather than infection control [7]. While multiple conventions exist to describe ventilation rate (e.g., total volumetric flow, volumetric flow per person and area, outdoor air ventilation rates), the air exchange rate (ACR) is frequently used in health care settings and is commonly expressed in units of air changes per hour (ACH). The existing minimum standards for ACH vary based on building type. For example, according to ASHRAE, the minimum required total ACH in residential homes is 0.35 ACH, whereas schools should be designed for approximately ten times higher rates. However, most schools do not meet this standard, in practice [8]. The suggestion of increasing the target to 4 to 6 ACH is more consistent with rates set in hospitals, where higher ACH rates provide airborne infection control among patients, staff and visitors. There is diverse and substantial evidence that elevated ACH levels should be adopted in routine indoor environments throughout society to reduce the spread of respiratory infections. A 2022 Italian schools study [9] found that effective ventilation systems can reduce the transmission of COVID-19 in schools by greater than 80%. This study, supported by the David Hume Foundation, compared coronavirus transmission clusters in 10,441 classrooms in Italy’s central Marche region. COVID clusters were found to be significantly lower in the 316 classrooms with mechanical ventilation systems, with the reduction level determined to the strength of the ventilation system (i.e., the transmission was inversely related to ACH level).

In general, the air changes per hour (ACH) are calculated by dividing the cubic feet of air per hour moved by the volume of space. This volumetric flow rate is often determined using a blower test to measure the cubic feet per minute moved and then multiplying that rate by 60 (minutes). However, this method does not account for in-room air mixing factors and provides limited information on ventilation efficiency within the room (e.g., the effectiveness of ventilation for the removal of a particular aerosol particle size range or contaminant type). Previous studies determined effective ventilation rates using tracer gasses such as sulfur hexafluoride (SF_6_) and carbon dioxide (CO_2_). While these perform well, they cannot be used with occupants in the room (confounding by occupant-generated CO_2_), have environmental concerns associated with their use (SF_6_), and cannot directly measure the impact of aerosol removal through filtration - which is the most common and cost-effective method for providing clean air to a space (via HVAC filters and in-room HEPA purifiers, for example).

Airflow-based determinations of ACH do not account for the full range of aerosol removal processes (filtration, deposition, etc), however understanding the effective aerosol removal rate is critical to providing airborne infection control. Effective air change rate (eACH) is a more representative metric that uses direct empirical measurements of aerosol particle removal in a space, rather than calculated estimates of total air exchange. Effective air change rate measurements accurately account for non-well mixed (also known as poorly mixed or stratified) conditions and all particle removal mechanisms. For this reason it is a better metric for infection control of pathogens that are carried by respiratory aerosol emissions. Further, since eACH measurements are taken within the breathing zone zone (defined by ASHRAE as between 3-72 inches) it is also a better measure of the ventilation building occupants are exposed to. Effective ACH can also be understood as a direct way to measure the related metric of equivalent air changes per hour (ACHe) added from outdoor air (which is calculated as the sum of non-infectious air delivery mechanisms that are active for a given room) but is more comprehensive as it includes passive aerosol removal mechanisms and takes into account natural ventilation.

Tracer aerosol technologies offer a practical method to directly measure the eACH of a space. For example, water-based DNA tracer particles offer stability, can be used in occupied spaces due to their harmless nature, and are subject to all aerosol removal mechanisms, along with filtration. These tracers offer direct measurement of pathogen transport and are an improved means of measuring the removal rates of liquid aerosols, such as respiratory aerosols, which may harbor pathogens. In a recent study [10], Arvelo et al. compared DNA-tagged aerosol decay rates to SF_6_ and CO_2_ decay rates. They demonstrated that DNA tracer aerosols can measure air change rates under well mixed conditions, and the effectiveness of aerosol control strategies such as HVAC filter grades can be evaluated by monitoring the decay rate of DNA tracer particle concentrations.

In this study, we demonstrate that DNA tracer particles combined with real-time sensors can be used to make rapid, accurate measurements of the effective air change rate (eACH) for respiratory aerosols within the breathing zone of non-well mixed rooms.

We examine the relationship between this approach, which estimates the rate of airborne pathogen removal between occupants in a room, and traditional ACH values derived from airflow measurements taken at HVAC supply vents. We also examine the effect of the level of air mixing (i.e., homogeneity or heterogeneity of particle distribution within a room) and decay window selection (i.e., measurement time).

We use two experimental test beds for these analyses. First, numerical simulation (computational fluid dynamic simulation, CFD) is conducted to visualize airflow and particle removal paths within a realistic large room. Here, simulated sensors are placed in concentric zones around a nebulizer providing test-particle releases, measuring the particle concentration every 15 seconds. This CFD model allows a direct comparison of the differences between eACH and airflow ACH values under varying levels of mixing and airflow, in a fully controlled system.

We then recapitulate this system in physical space to validate the CFD results under real-world conditions that include all mechanisms of particle removal that contribute to true aerosol clearance rates, including deposition and leakage. We measure eACH using the decay of DNA tracer aerosols nebulized and monitored in real-time by Poppy Sensors. Airflow and particle motion in the room is induced by forced mechanical ventilation and other mechanisms. Finally, we find that a standard sampling time of 15 minutes from the end of nebulization is sufficient to produce an accurate eACH value under non-well mixed conditions.

## 2. Test Space (physical and virtual)

In indoor environments, airflow is either achieved by structural portals (doors, windows, etc.) or through a mechanical feed of air through ventilation systems. Different airflow scenarios can significantly influence particle movement, as numerous studies have shown [11, 12, 13]. For example, one numerical study showed that particles exhaled by a person can remain suspended in the air for several minutes in a supermarket aisle [14]. Another study showed that opening the window in a classroom near an infected teacher allowed particles to escape via the window and considerably reduced particle movement across the room [15]. As the layout of every commercial building differs, it is crucial to minimize the number of domain-related variables that potentially influence particles from spreading differently. To simulate a real-world scenario, unlike most studies which are conducted in small environments, this study was conducted in a large test space (Figure 1) (ARE Labs, 15320 S. Cornice St., Olathe, KS-66062), which is representative of a typical open-plan office. This space was also modeled closely in our CFD analyses. The room has an area of 1673 ft^2^ with a ceiling height of 15 ft., which corresponds to a room volume of 25095 ft^3^. See **Appendix B** for detailed dimensions of the test space.

**Figure 1.**
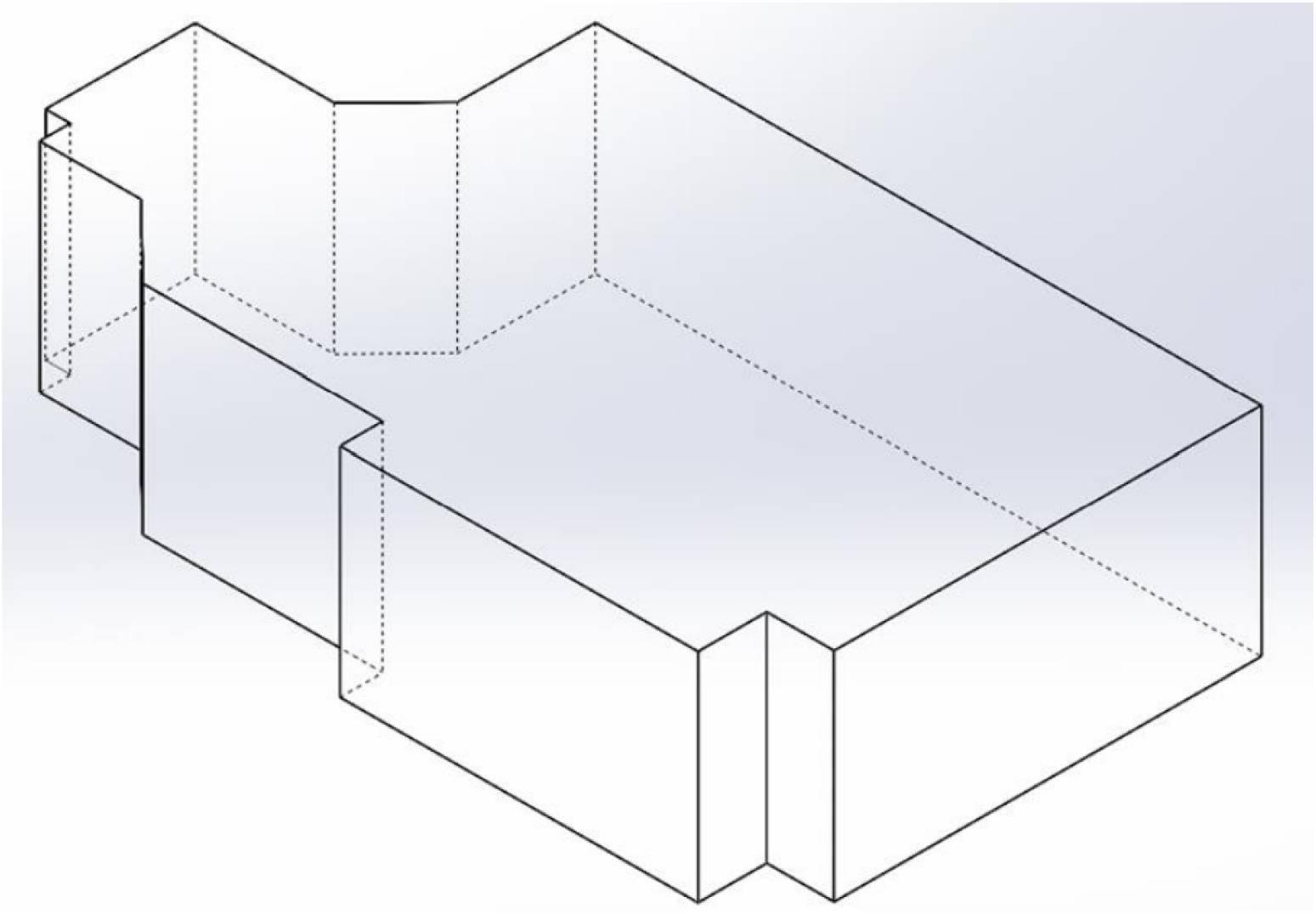
Simplified 3D CAD version of the test room. The room has an area of 1673 ft^2^ with a ceiling height of 15 ft., which corresponds to a room volume of 25095 ft^3^.

## 3. Numerical Model - Computational Fluid Dynamics

A numerical study was performed to model the natural distribution of air and the reduction of airborne particles with airflow inside the large indoor test space. Furthermore, particle decay computed at simulated sensor locations around a source in the test space from the numerical study is compared with experimental results attained using Poppy Sensors inside the physical version of the same space.

Computational Fluid Dynamics is a subset of Fluid Mechanics, where the equations of flow (Navier-Stokes) are solved numerically using computation. The geometry is divided into smaller elements, and the equations of fluid motion are solved at every element for a specified time duration to obtain an accurate estimation of the fluid flow and particles which travel with the flow. The flow equations are numerically discretized and applied to the meshed geometries to calculate the change in the physical properties after each iteration. CFD can be used as a validation tool to compare the experimental results with the theoretically modeled results.

### 3.1 General Steps

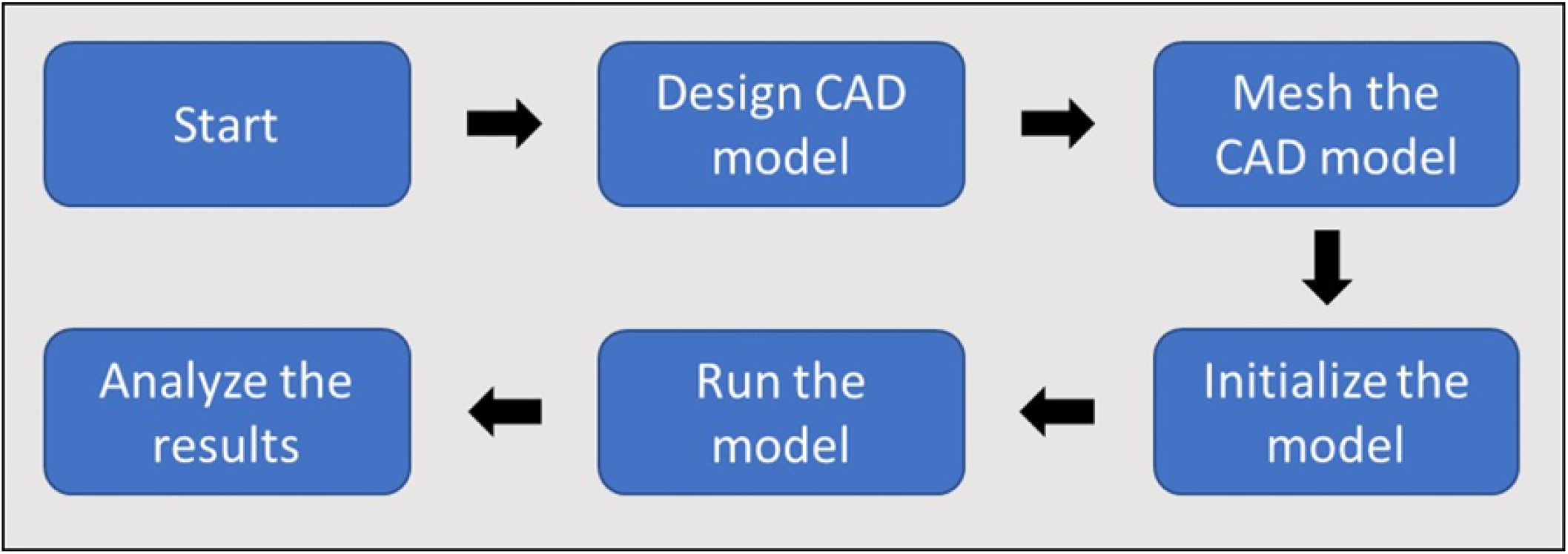

The CFD analysis can be summarized from the following sequence:

1. <u>Design of CAD model</u>: The numerical model is first visualized and then designed using CAD software. This step aims to recreate the physical operating environment of the experimental trials, draft a theoretical scenario for the device operation, and generate an accurate 3D/2D model that best describes the space.

2. <u>Mesh the CAD model</u>: The CAD model design is meshed so that numerical runs can be performed on them. Meshing is the technique in which geometry is divided into smaller bits or grids to solve the test variables using the data inside these grids. Meshing can be done in two ways, finite element meshing and finite volume meshing. In finite elements, the solid object is divided into grids, and calculations are done at each grid point. In finite-volume meshing, the body is divided into smaller volumes. This method is preferred for the CFD analysis as the conservation principles (mass, momentum, and energy transport) can be satisfied. These conditions drive the model.

3. <u>Initialize the model</u>: Initializing a CFD model involves setting the boundary conditions, discretizing the operating equations based on the model, and providing realistic guess values to the model to start the numerical simulation. Boundary conditions are the settings applied to faces or the boundaries inside the designed model. These may be the fluid boundary conditions (velocity inlet, pressure outlet, etc.) and the wall boundary conditions (wall slip, no-slip, etc.). These conditions play an essential role in achieving desired results. Discretization is a numerical method of breaking down partial differential equations (PDEs) and ordinary differential equations (ODEs) into node-based equations using Taylor Series Expansion. Discretization enables the study to control the order of magnitude for the errors based on the governing equations and type of study. For running any numerical study, a guess value (start value) must be provided to the model. The value provided must be realistic and within the permitted limits of the model, as providing non-realistic values can generate inconsistent results, which might be expensive computationally.

4. <u>Run the model</u>: Using the guess values provided during the initialization, the model is solved for the test variables until a specific condition is met, which can either be the relative error or a set number of time steps.

### 3.2 ANSYS Fluent

ANSYS Fluent is a full-featured fluid dynamics solution for modeling flow and other related physical phenomena that offers unparalleled analysis capabilities. It provides all the tools needed to design and optimize new equipment and troubleshoot existing installations. The versatile technology offers insight into how a product design will behave in the real world, all before a single prototype is built. It includes solvers that accurately simulate the behavior of the broad range of flows that engineers encounter daily — from Newtonian to non-Newtonian, from single-phase to multi-phase, and from subsonic to hypersonic. It also can solve highly complicated multiphysics problems with high accuracy and speed. The analysis performed for this project was performed using the Ansys FLUENT 2021R2

### 3.3 Test Room Design and Room-Specific Flow Conditions

The HVAC ventilation inside a room is governed by the American Society of Heating Ventilation and Air Conditioning Engineers (ASHRAE) standard, which standardizes the minimum required flow rate inside a space based on the number of people operating inside the room, type of the room (residential [ASHRAE standard 62.2] or commercial [ASHRAE standard 62.1]), the volume of the room, etc. The current room geometry was taken, and the existing HVAC design code was employed to design an HVAC system to simulate airflow and particle decay. The vent design and placement inside a room is governed by the Sheet Metal and Air Conditioning Contractors’ National Association (SMACNA). As per ASHRAE, the cooling capacity of the HVAC unit to cool this room is estimated to be around 5t (5 Ton). The minimum flow rate to operate a 5t HVAC system is 2000 CFM. Therefore, the total flow rate inside the modeled room for this study was 2000 CFM.

#### 3.3.1 Case I: The Standard HVAC Layout Case

Based on the standards stated previously, the flow rate inside the room was set to 2000 CFM. The room was modeled to have ten supply and ten return units, each operating at 200 CFM. The design of the room with ten supply and ten return units can be seen in Figure 2. This study was performed to achieve predictions for two conditions: non-well mixed and well mixed. The *non-well mixed condition* refers to a non-uniform distribution of particles across the volume. It is simulated by placing mixing fans inside the chamber to introduce recirculating zones so that particles that uniformly spread in the volume at the start of the simulation are disturbed. In *well-mixed conditions*, the simulation was performed with no mixing fans inside the test space, i.e. the simulation starts with uniformly distributed particles in the whole volume of the space. The table below describes the initial and boundary conditions used for this case.

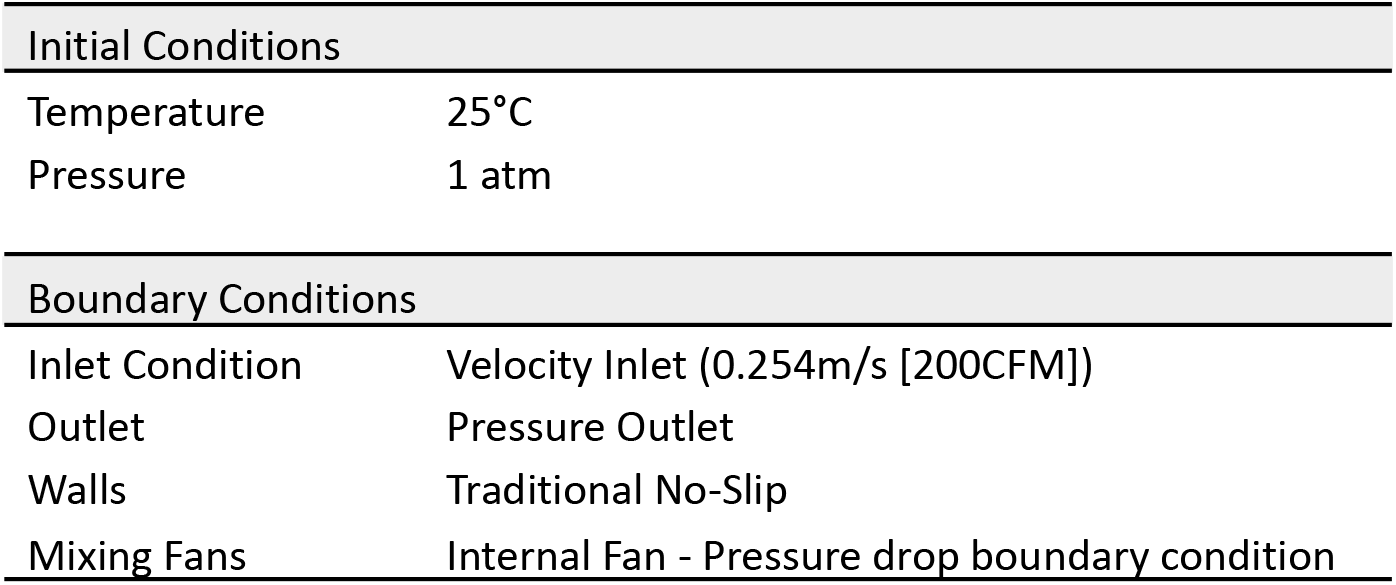

**Figure 2.**
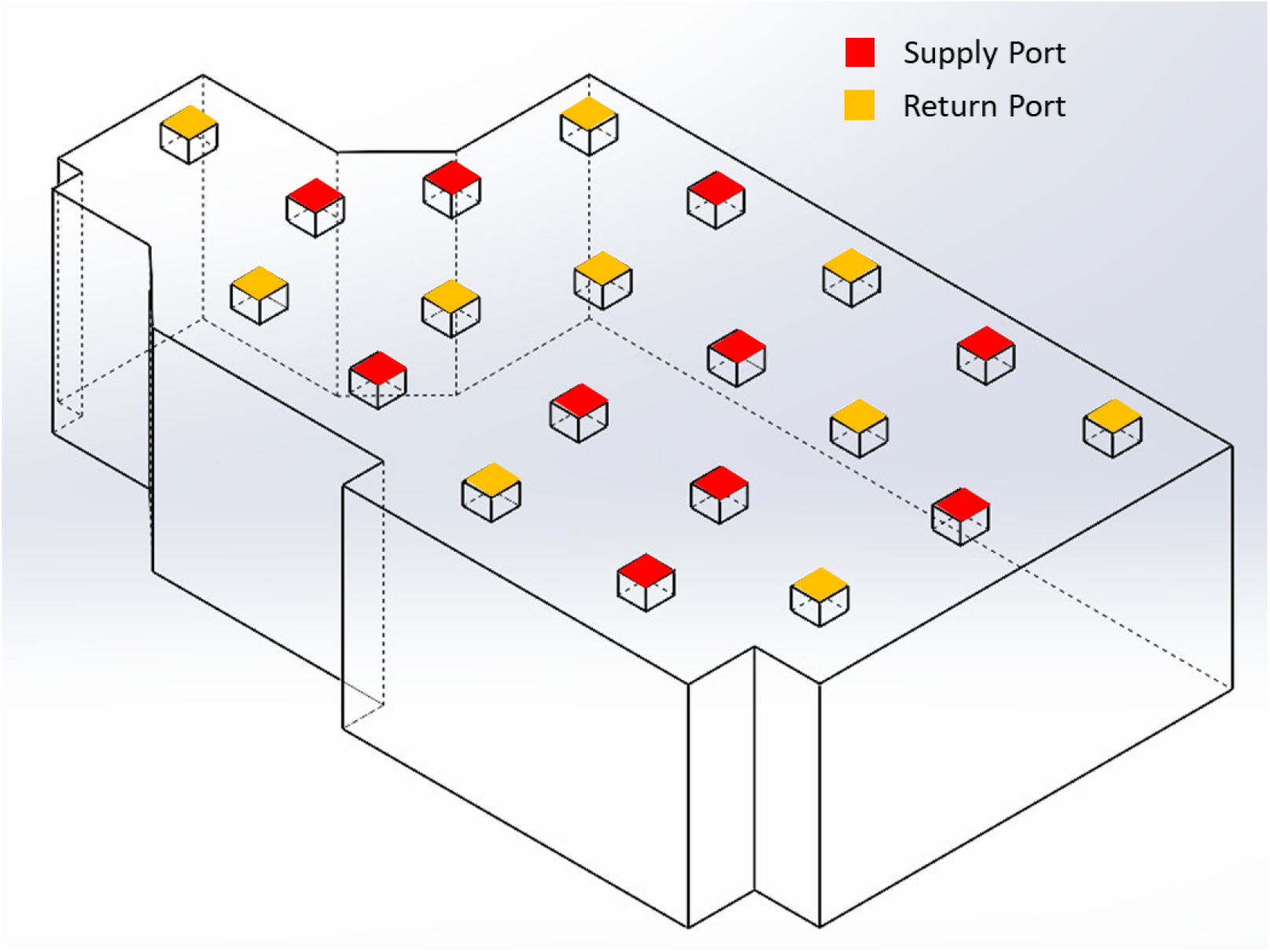
The geometric setup of the standard HVAC case with 10 supply units and 10 return units designed according to the ASHRAE and SMACNA Standards.

It is important to note that mixing fans are used in CFD modeling to create a non-well mixed condition as the CFD software generates a uniform distribution of particles (well mixed condition). The non-well mixed condition uses mixing fans to disturb this uniformity. In contrast, later in the study when realistic and physical tests are done within a live test space, mixing fans are used in order to generate a uniform distribution of particles. Therefore, mixing fans are used to create a wellmixed condition for live experiments, but they generate a non-well mixed condition in CFD modeling.

The standard HVAC design numerical study was conducted in two steps:

1. Obtaining the steady state solution for the flow inside the test chamber; a steady state solution implies that the flow inside the chamber was solved until a constant flow pattern was obtained.
2. Based on the steady state flow patterns, a particle tracking operation was performed where 1 million particles were introduced into the chamber with the assumption that they would flow along the already established fluid flow inside the chamber and would not be collected on the chamber floor. The properties of the particles can be seen in Appendix A.

##### Well Mixed Condition - Uniform Spread of Particles

###### Steady State Solution

A turbulent k-ε model was set up using ANSYS Fluent, with the air coming out of each supply duct set at 50 ft/min or 0.254 m/s. The walls were set to the no-slip boundary condition (which meant that the velocity at the walls was zero). The model had 10 return units, which were set as pressure outlets to balance the system. The flow inside the chamber was validated to be a constant 2000 CFM. The geometric setup of the modeled space can be seen in Figure 2. Watertight geometry workflow was used to perform the meshing of the CFD analysis. The standard local sizing and surface meshing settings were used to generate local, and surface meshes. Boundary layers were added at the walls of the geometry for accurate flow calculations at the walls, and a poly hexacore volumetric meshing was used to generate the computational cells. The meshing process generated an average of 1 million computational cells.

The simulation was run until the solution satisfied a convergence criterion set as 1 x e^-3^. Convergence is the state in a numerical simulation where the relative errors for the test variables reduce to a set threshold. Every fluid flow simulation has four minimum convergence criteria: continuity, x-momentum, y-momentum, z-momentum. Since we used a turbulent k-e model for simulation, we had two extra convergence criteria, k and e. The solution is said to be fully converged when the residuals (error values) values between iterations go below a set value. For the simulation in this CFD simulation, it was set to 1e-3. This was applied to continuity, x-momentum, y-momentum, z-momentum. The k and e convergence criterias were set to 1e-6.

After getting a converged solution, the flow inside the chamber can be seen in Figure 3. It was observed that the air enters the chamber from the inlet ducts and moves straight down toward the floor. As it collides with the floor, it diffuses into the adjacent areas of the chamber until it finally starts to exit the chamber. Due to the inlet duct placement, the flow field allowed every corner of the chamber to be filled with air, ensuring maximum airflow coverage. From visual validation, the flow inside the chamber was as expected, and this solution was used as the set flow pattern for the particle tracking analysis.

**Figure 3.**
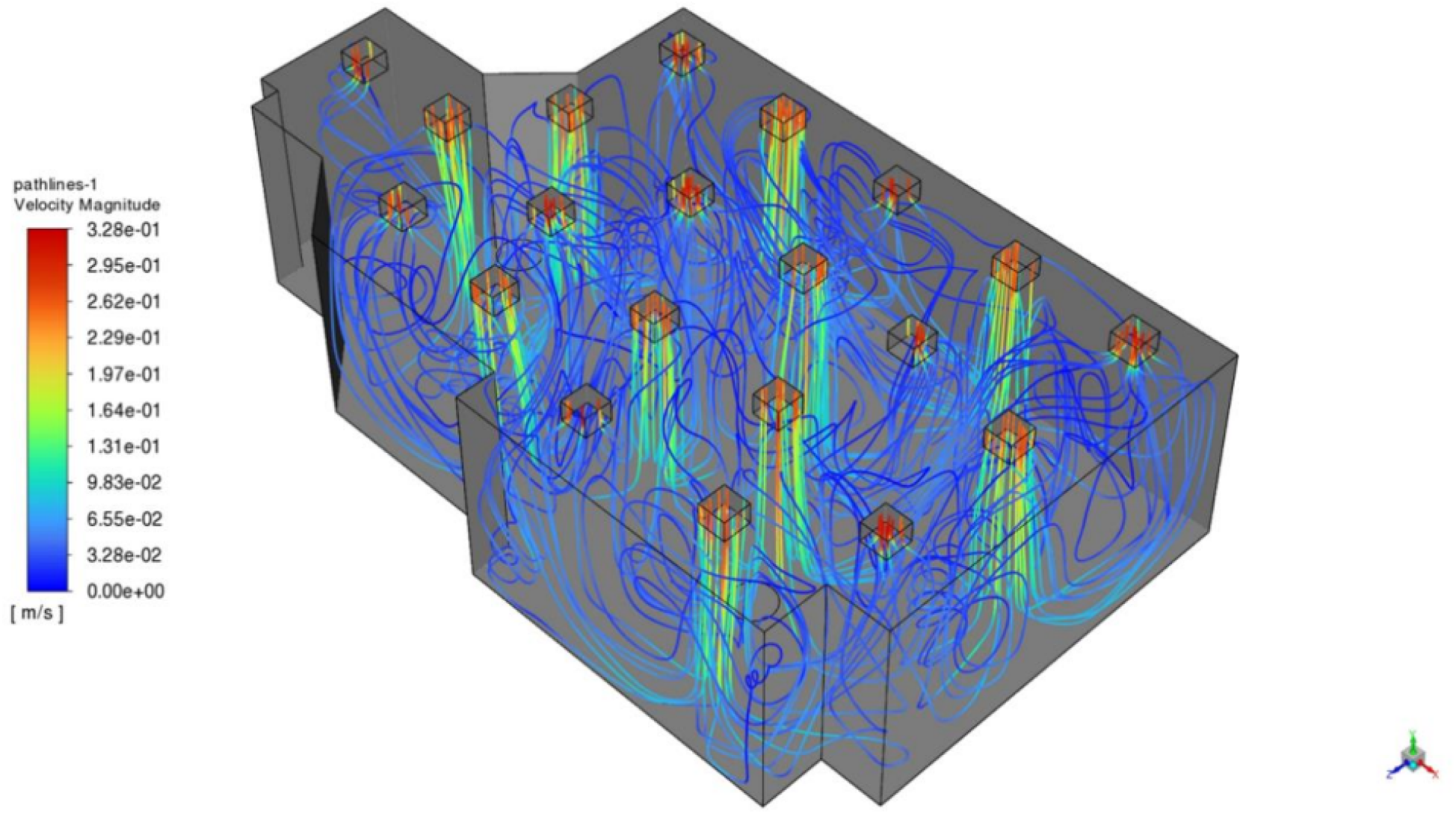
Flow inside the standard HVAC layout, numerically simulated chamber with 10 supply vents and 10 return vents, being operated to achieve a total room flow rate of 2000 CFM.

###### Particle Tracking Analysis

After confirming the results, a transient Discrete Phase Model (DPM) was employed onto the steady state solution. The particles inside the chamber were introduced uniformly in the first-time step of the study, and the total simulation was performed for 120 computational minutes. The reduction in the concentration of the particles inside the chamber during the total simulation can be seen in Figure 4. It was observed that the particles inside the chamber moved along the airflow and started exiting through the chamber’s return ducts. From the graph, the natural particle reduction inside the chamber due to the flow field achieved a 3.37 Log reduction in 120 minutes, tested numerically.

**Figure 4.**
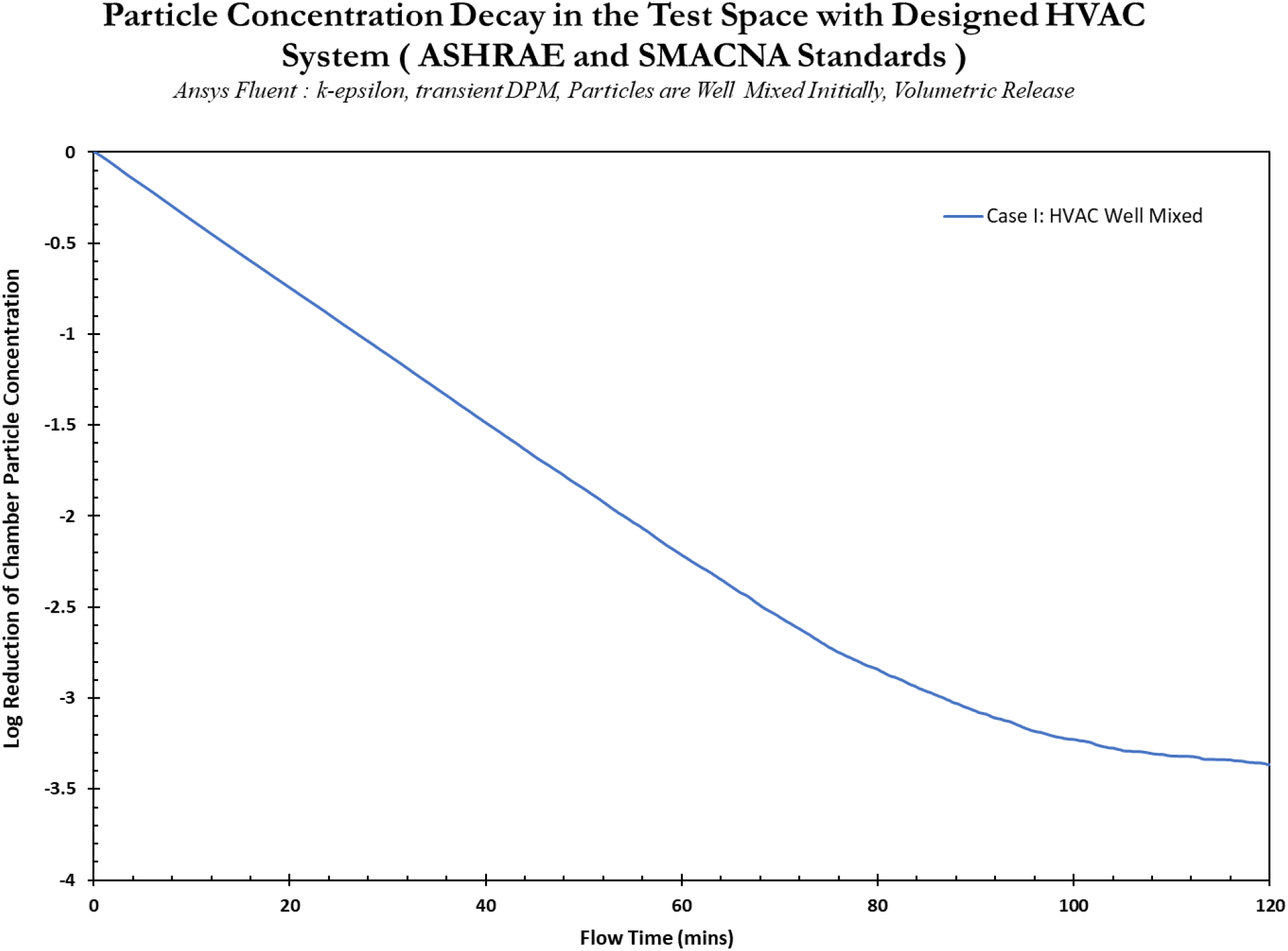
Total reduction of particles inside the standard HVAC test chamber, with 10 supply vents and 10 return vents, operated to achieve a total room flow rate of 2000 CFM.

##### Non-Well Mixed Condition - Mixing Fans to Disrupt Uniform Spread of Particles

###### Steady State Solution

Another turbulent k-ε model was set up using ANSYS Fluent, with the air coming out of each supply duct set at 50 ft/min or 0.254 m/s. The walls were set to the no-slip boundary condition (which meant that the velocity at the walls was zero). The model had 10 return units, which were set as pressure outlets to balance the system. The flow inside the chamber was validated to be a constant 2000 CFM. Two mixing fans were introduced into the room to help the mixing inside the chamber. Internal fan boundary condition was used to simulate these fans’ operation, and they were set to operate at 750 CFM. The geometric setup of the modeled space can be seen in Figure 5. Watertight geometry workflow was used to perform the meshing of the CFD analysis. The standard local sizing and surface meshing settings were used to generate local, and surface meshes. Boundary layers were added at the walls of the geometry for accurate flow calculations at the walls, and a poly hexacore volumetric meshing was used to generate the computational cells. The meshing process generated an average of 1 million computational cells.

**Figure 5.**
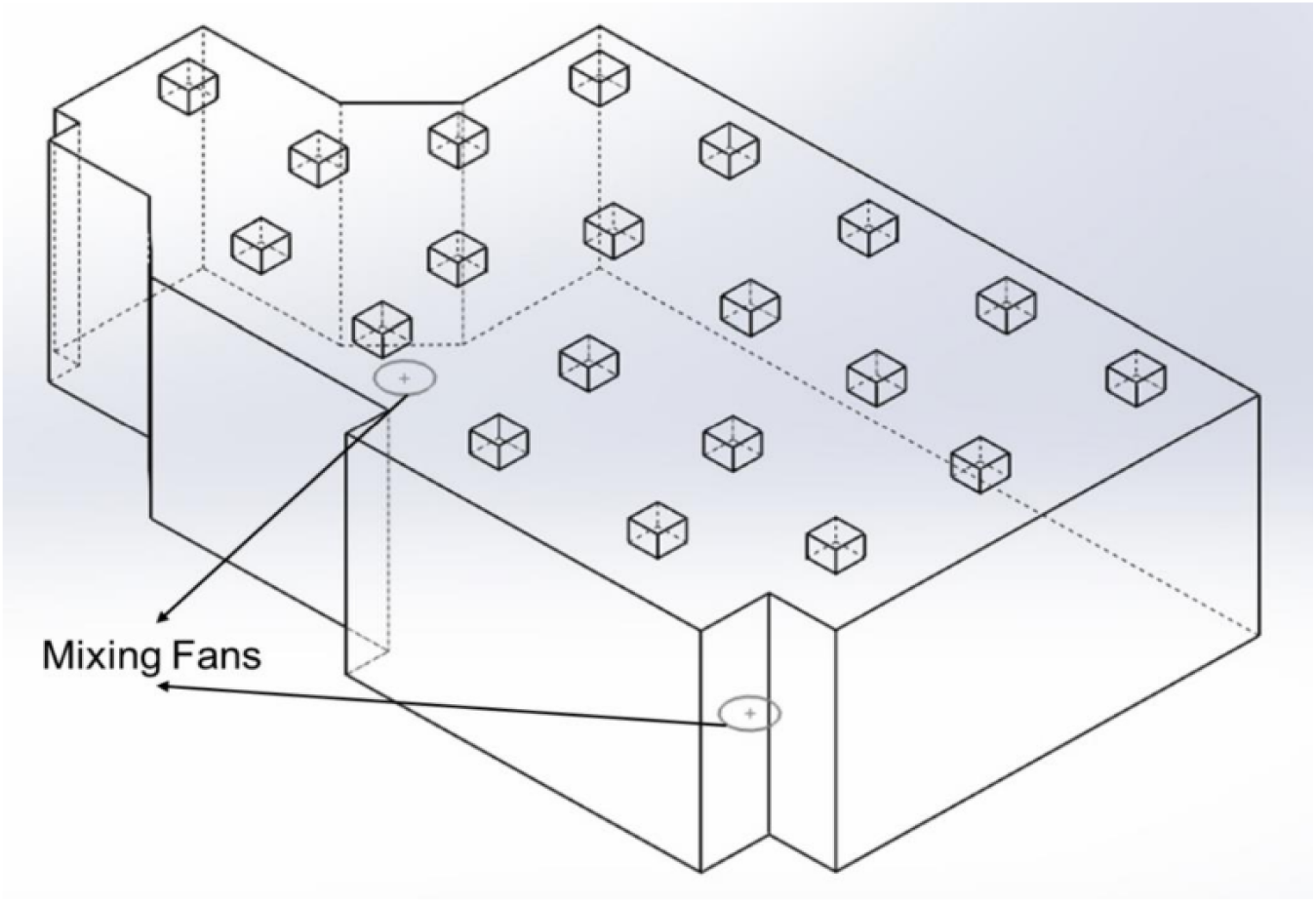
The geometric setup of the standard HVAC case with 10 supply units and 10 return units designed according to the ASHRAE and SMACNA Standards, with mixing fans placed inside the room.

The simulation was run until the solution satisfied a convergence criterion. The flow inside the chamber after getting a converged solution can be seen in Figure 6. From Figure 6, it was observed that the air enters the chamber from the inlet ducts, moving straight down toward the floor. Then, as it collides with the floor, it diffuses into the chamber’s adjacent areas until it finally starts to exit.

**Figure 6.**
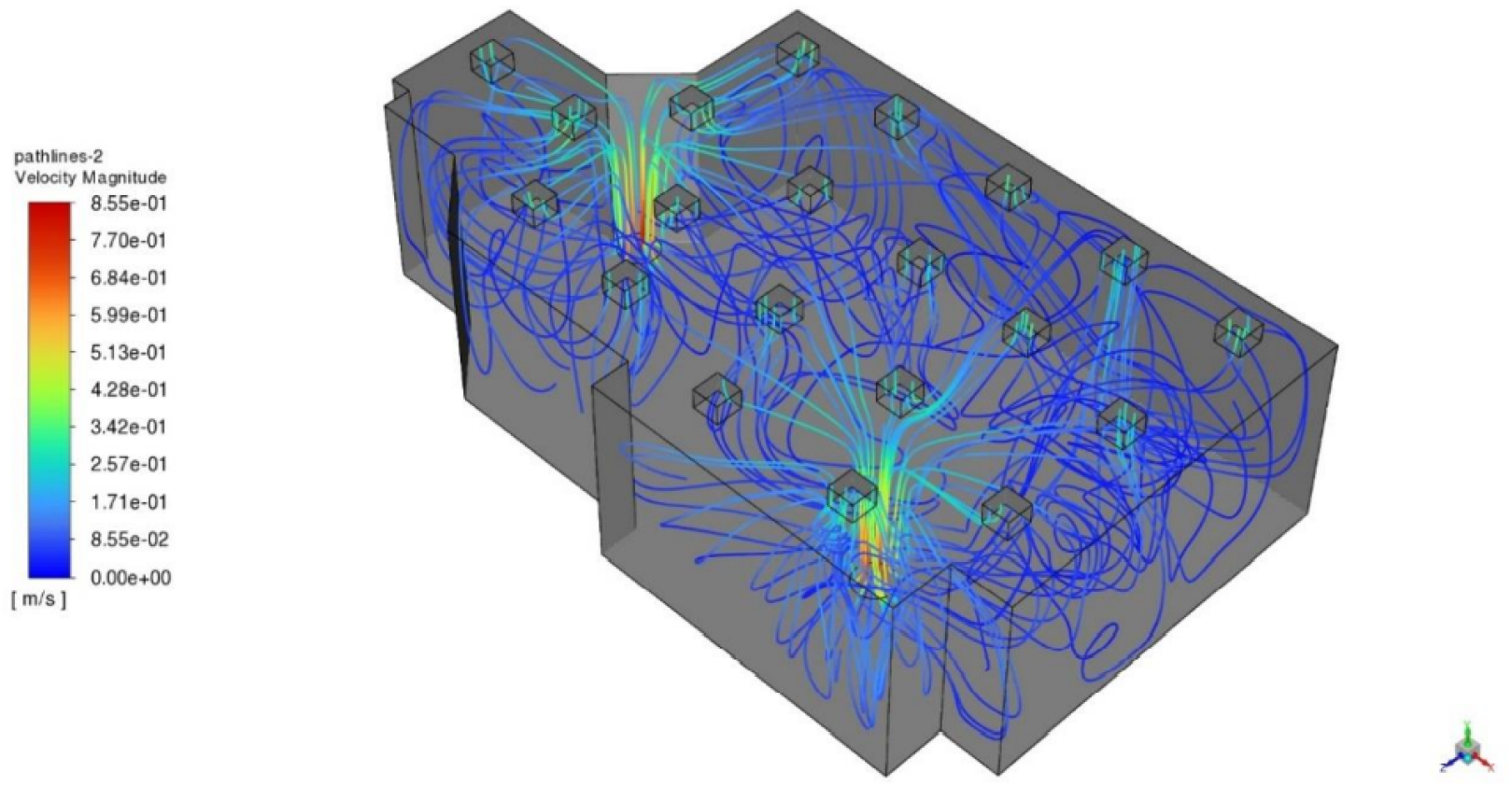
Flow inside the standard HVAC layout, numerically simulated chamber with 10 supply vents and 10 return vents, being operated to achieve a total room flow rate of 2000 CFM, with mixing fans recirculating the air inside the chamber.

Due to the inlet duct placement, the flow field allowed every corner of the chamber to be filled with air, ensuring maximum airflow coverage. The mixing fans introduce local recirculation zones and mix the particles causing small pockets of non-uniformity. From visual validation, the flow inside the chamber was as expected, and this solution was used as the set flow pattern for the particle tracking analysis.

###### Particle Tracking Analysis

After confirming the results, a transient Discrete Phase Model (DPM) was employed onto the steady state solution. The particles inside the chamber were introduced uniformly in the first time-step of the study, and the total simulation was performed for 120 computational minutes. The reduction in the concentration of the particles inside the chamber during the entire simulation can be seen from Figure 7.

**Figure 7.**
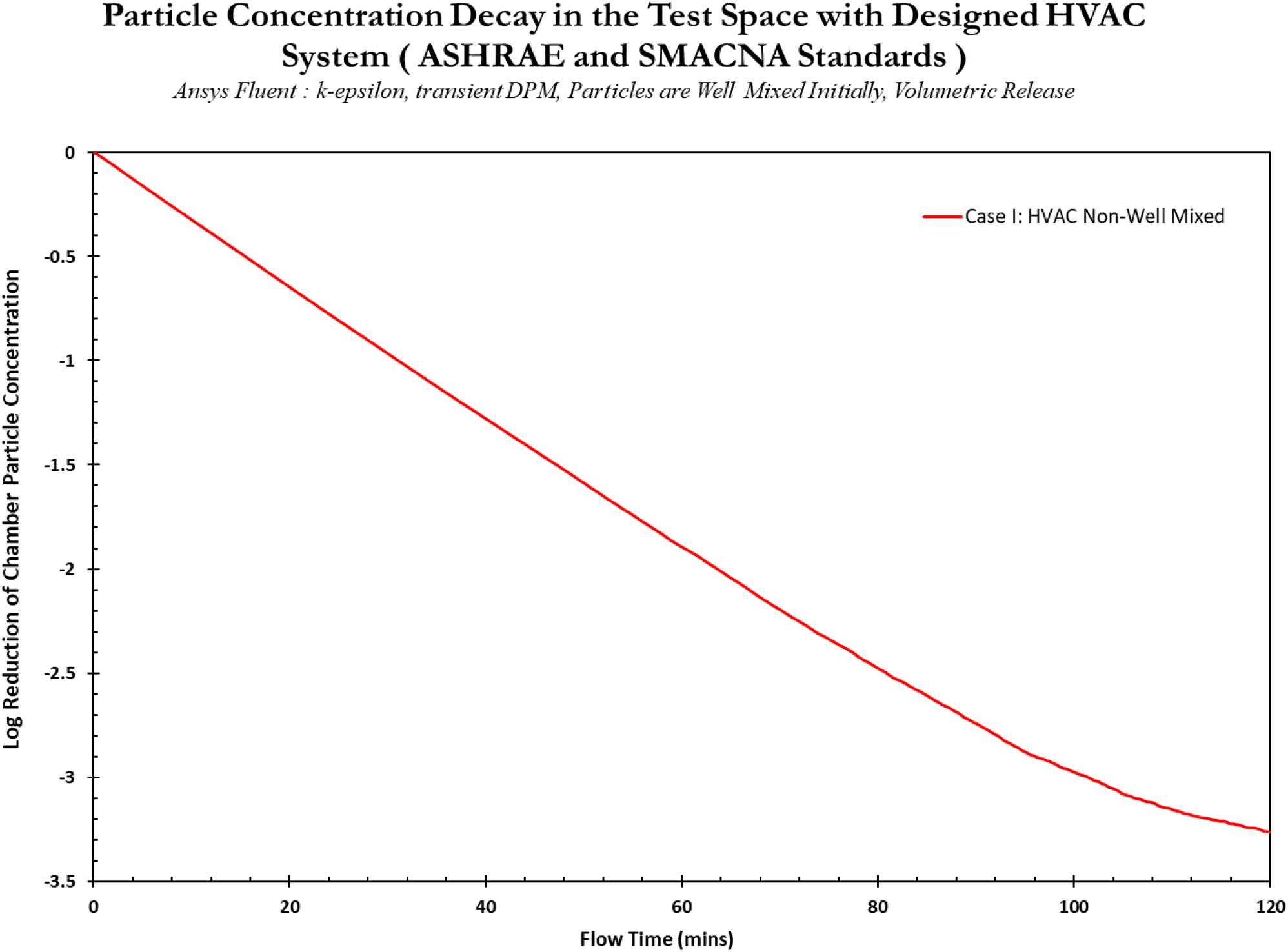
Total reduction of particles inside the standard HVAC test chamber (with ducts and supply vents designed to ASHRAE and SMACNA standards), with 10 supply vents and 10 return vents, being operated to achieve a total room flow rate of 2000 CFM, with mixing fans recirculating the air inside the chamber.

It was observed that the particles inside the chamber moved along the airflow and started exiting through the chamber’s return ducts. Due to the flow field, the natural particle reduction inside the chamber achieved a 3.27 Log reduction in 120 minutes of numerical testing (Figure 7).

###### Brief Conclusion

By performing the above analysis, we observed that the reduction inside the chamber for the well mixed case was faster than the non-well mixed case (Figure 8). This observation was expected, as with uniform loading when particles are spread out evenly they exit the chamber faster than when non-uniformities are present (i.e. in this case, when mixing fans are introduced). The recirculation introduced by the mixing fans increased the residence time of the particles, which were already very well mixed based on the uniform particle distribution at simulation starting time, and by virtue of vent placement, creating minor deviations in their path while exiting the chamber.

**Figure 8.**
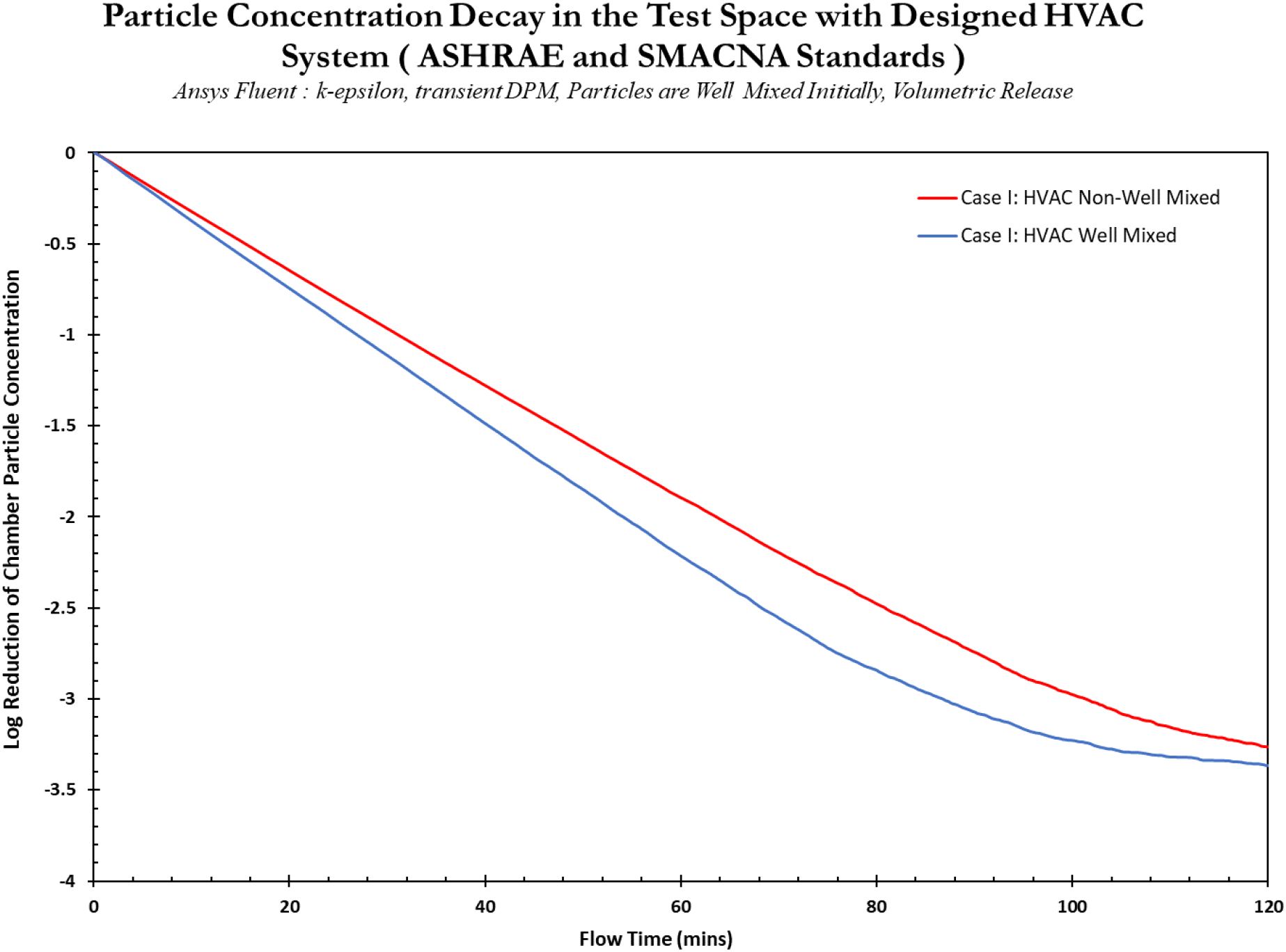
Total reduction of particles inside the virtual test chamber, with 10 supply vents and 10 return vents, being operated to achieve a total room flow rate of 2000 CFM; Comparing standard HVAC case without and with mixing fans

#### 3.3.2 Case II: The Designed Ventilation System Case

To create an environment capable of airflow patterns that can better help us explore the effects of real world non-well mixed conditions on eACH measurements, we designed a ventilation system that can be used to drive multiple flow conditions. The system has one inlet pipe and one outlet pipe placed strategically within the test space (Figure 9). In addition, the room was set up to achieve a realistic arrangement of components so that these simulations could be replicated in physical testing. The table below describes the initial and boundary conditions used for this case.

| Initial Conditions |  |
| --- | --- |
| Temperature | 25°C |
| Pressure | 1 atm |
| Boundary Conditions |  |
| Inlet Condition | Velocity Inlet (4.68m/s) |
| Outlet | Pressure Outlet |
| Walls | Traditional No-Slip |
| Mixing Fans | Internal Fan - Pressure drop boundary condition |

**Figure 9.**
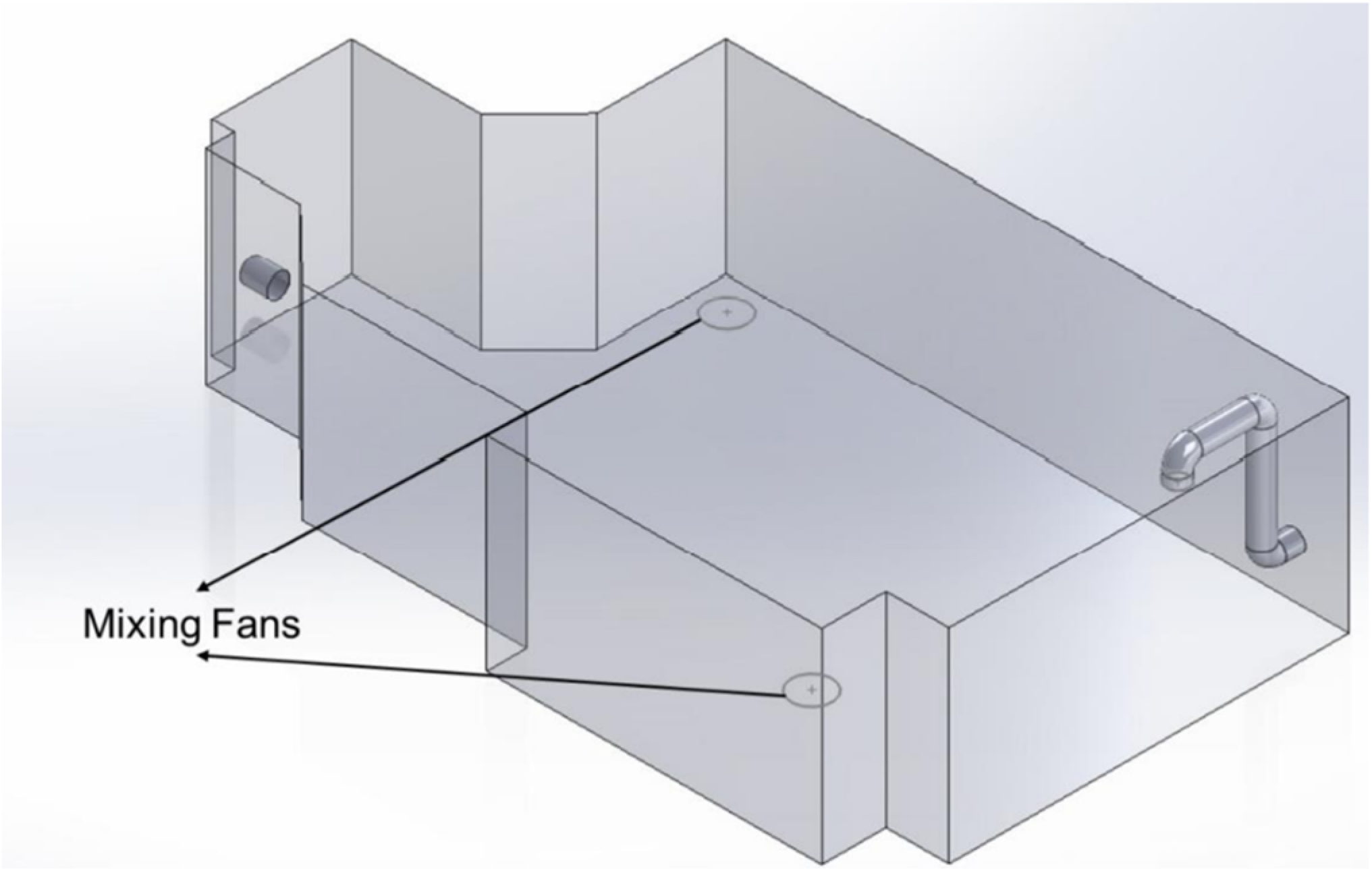
The geometric setup of the designed ventilation system case with one inlet pipe and one outlet pipe inside the room. The airflow inside the room was designed to operate at 2000 CFM.

This study was performed in well mixed and non-well mixed scenarios to investigate if particle decay in the test space is similar to the results for the standard HVAC system design. Here, the mixing fans were employed to maintain homogeneity for a well-mixed case, while the default configuration of volumetric loading is a non-well mixed condition.

The numerical study was conducted in two steps.

1. Obtaining the steady state solution for the flow inside the test chamber; a steady state solution implies that the flow inside the chamber was solved until a constant flow pattern was obtained.
2. Based on the steady state flow patterns, a particle tracking operation was performed where 1 million particles were introduced into the chamber with the assumption that they would flow along the already established fluid flow inside the chamber and would not be collected on the chamber floor. The properties of the particles can be seen in Appendix A.

##### Well Mixed Condition – Mixing Fans to Maintain Homogeneity of Particles

###### Steady State Solution

A turbulent k-ε model was set up using ANSYS Fluent with the air coming out of the inlet pipe, whose diameter was 20 inches, with a flow velocity of 916.37 ft/min or 4.68 m/s. The walls were set to the no-slip boundary condition (which meant that the velocity at the walls was zero). The model had an outlet pipe with an outer diameter of 20 inches, which was set as a pressure outlet to balance the system. The flow inside the chamber was a constant 2000 CFM. Two mixing fans were introduced into the room to help the mixing inside the chamber. Internal fan boundary condition was used to simulate these fans’ operation, and they were set to operate at 750 CFM. The geometric setup of the modeled space can be seen in Figure 9. Watertight geometry workflow was used to perform the meshing of the CFD analysis. The standard local sizing and surface meshing settings were used to generate local, and surface meshes. Boundary layers were added at the walls of the geometry for accurate flow calculations at the walls, and a poly hexacore volumetric meshing was used to generate the computational cells. The meshing process generated an average of 1 million computational cells.

The simulation was run until the solution satisfied a convergence criterion. The flow inside the chamber after reaching a converged solution can be seen in Figure 10. It was observed that the air enters the chamber from the inlet pipe, moving straight down toward the floor (Figure 10). It diffuses into the chamber’s adjacent areas as it collides with the floor. Since the inlet duct was situated in the corner of the chamber, the diffused air collided with the walls adjacent to the flow to create local vortex zones. The flow eventually exits its vortex and moves towards the outlet pipe to exit the chamber. From visual validation, the flow inside the chamber was as expected, and this solution was used as the set flow pattern for the particle tracking analysis.

**Figure 10.**
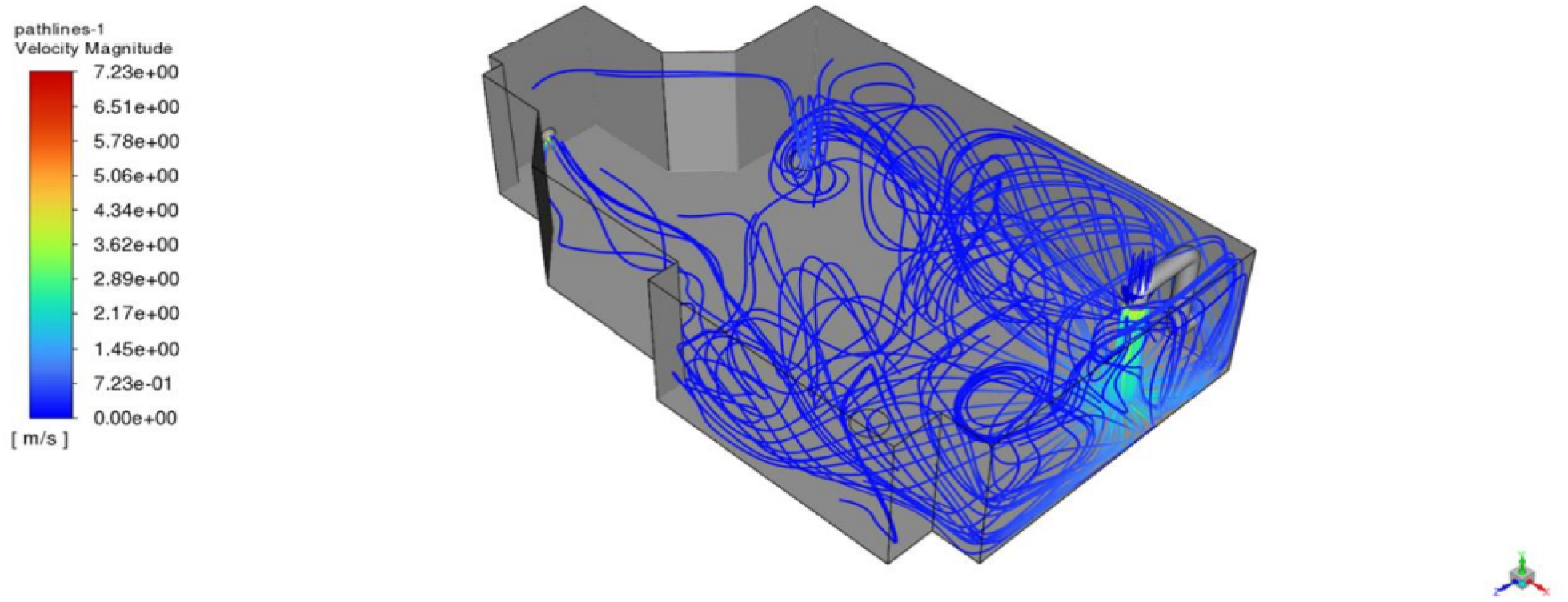
Flow inside the designed ventilation system, numerically simulated chamber with one air inlet and one air outlet inside the room. The airflow inside the room was designed to operate at 2000 CFM.

###### Particle Tracking Analysis

After confirming the results, a transient Discrete Phase Model (DPM) was employed onto the steady state solution. The particles inside the chamber were introduced uniformly in the first-time step of the study, and the total simulation was performed for 120 computational minutes. The reduction in the concentration of the particles inside the chamber during the total simulation can be seen from Figure 11. It was observed that the particles inside the chamber moved along the airflow and started exiting the chamber through the outlet pipe of the chamber. At the same time, the mixing fans were placed strategically inside the room to recirculate the local air around the fan to help the original flow from the inlet pipe to cover more area of the room. The natural particle reduction inside the chamber due to the flow field achieved a 2.37 Log reduction in 120 minutes, tested numerically (Figure 11).

**Figure 11.**
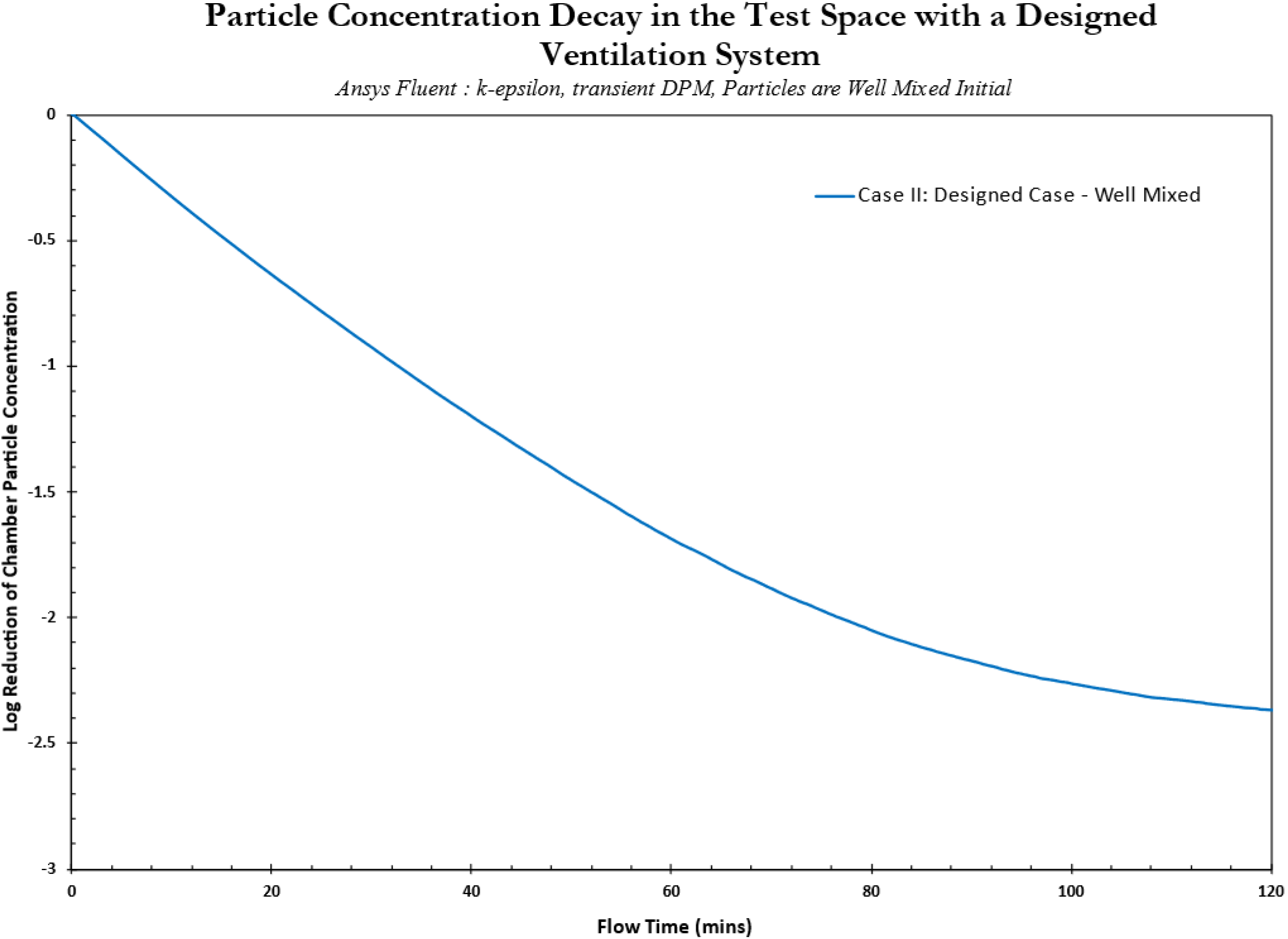
Total reduction of particles inside the designed ventilation system test chamber with one inlet pipe and one outlet pipe unit inside the room. The airflow inside the room was designed to operate at 2000 CFM.

##### Non-Well Mixed Condition - Mixing Fans to Disrupt Uniform Spread of Particles

###### Steady State Solution

A turbulent k-ε model was set up using ANSYS Fluent with the air coming out of the inlet pipe, whose diameter was 20 inches, with a flow velocity of 916.37 ft/min or 4.68 m/s. The walls were set to the no-slip boundary condition (which meant that the velocity at the walls was zero). The model had an outlet pipe with an outer diameter of 20 inches, which was set as a pressure outlet to balance the system. The flow inside the chamber was a constant 2000 CFM. The geometric setup of the modeled space can be seen in Figure 12.

**Figure 12.**
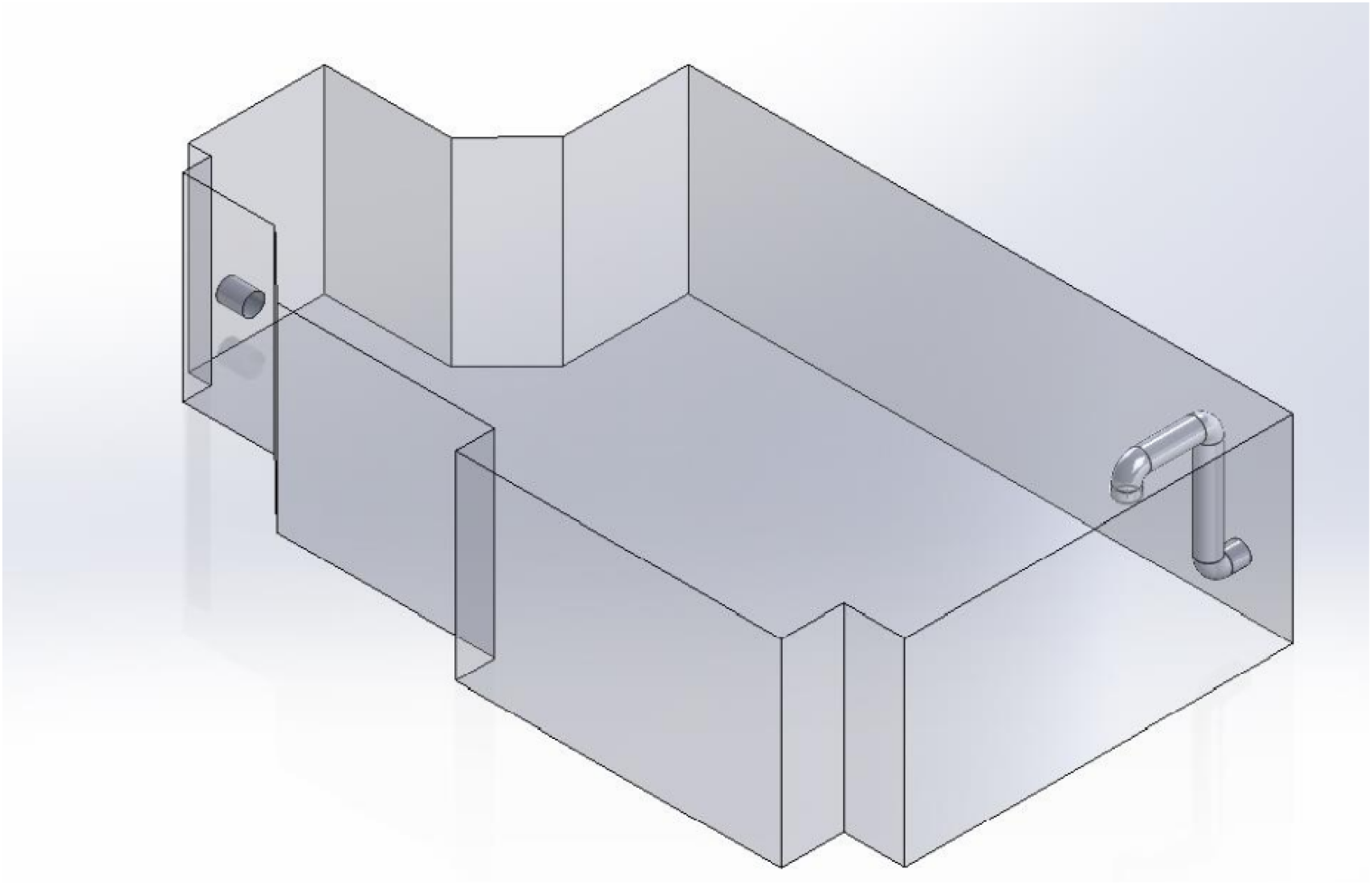
The geometric setup of the designed ventilation system case with one inlet pipe and one outlet pipe unit, with two mixing fans placed inside the room. The airflow inside the room was designed to operate at 2000 CFM.

Watertight geometry workflow was used to perform the meshing of the CFD analysis. The standard local sizing and surface meshing settings were used. Boundary layers were added at the walls of the geometry for accurate flow calculations, and a poly hexacore volumetric meshing. The meshing process generated an average of 1 million computational cells.

The simulation was run until the solution satisfied a convergence criterion. The flow inside the chamber after getting a converged solution can be seen in Figure 13.

**Figure 13.**
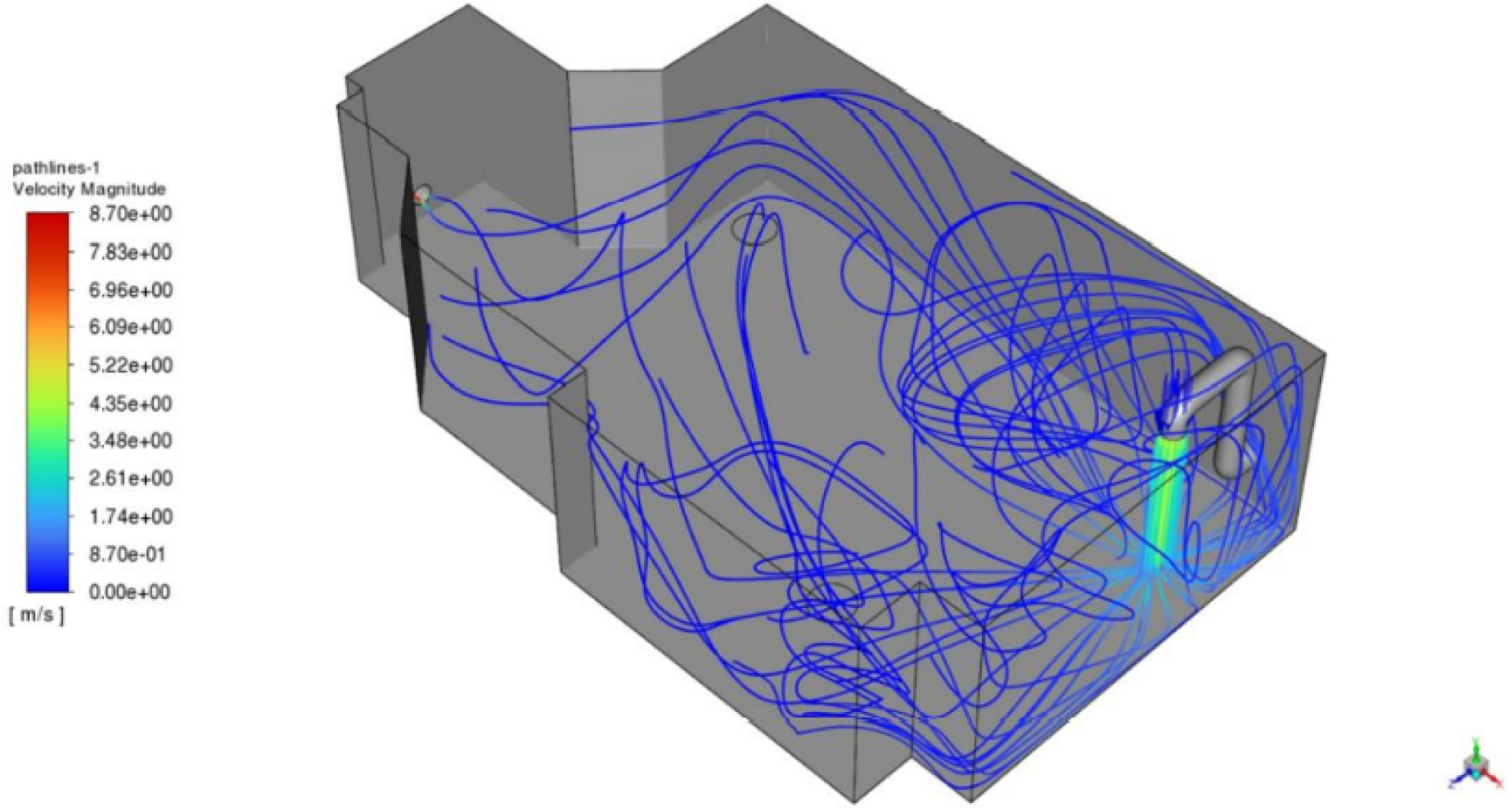
Flow inside the designed ventilation system, numerically simulated chamber with one inlet pipe and one outlet pipe unit, with mixing fans placed inside the room. The airflow inside the room was designed to operate at 2000 CFM.

It was observed that the air enters the chamber from the inlet pipe, moving straight down toward the floor (Figure 13). As it collides with the floor, it starts to diffuse into the chamber’s adjacent areas. Since the inlet duct was situated in the corner of the chamber, the diffused air collided with the walls adjacent to the flow to create local vortex zones. The flow eventually exits the chamber through the outlet pipe. From visual validation, the flow inside the chamber was as expected, and this solution was used as the set flow pattern for the particle tracking analysis.

###### Particle Tracking Analysis

After confirming the results, a transient DPM was employed onto the steady state solution. The particles inside the chamber were introduced uniformly in the first-time step of the study, and the total simulation was performed for 120 computational minutes. The reduction in the concentration of the particles inside the chamber during the total simulation can be seen in Figure 14. It was observed that the particles inside the chamber moved along the airflow and started exiting through the chamber’s return ducts. From the graph, the natural particle reduction inside the chamber due to the flow field achieved a 2.13 Log reduction in 120 minutes, tested numerically.

**Figure 14.**
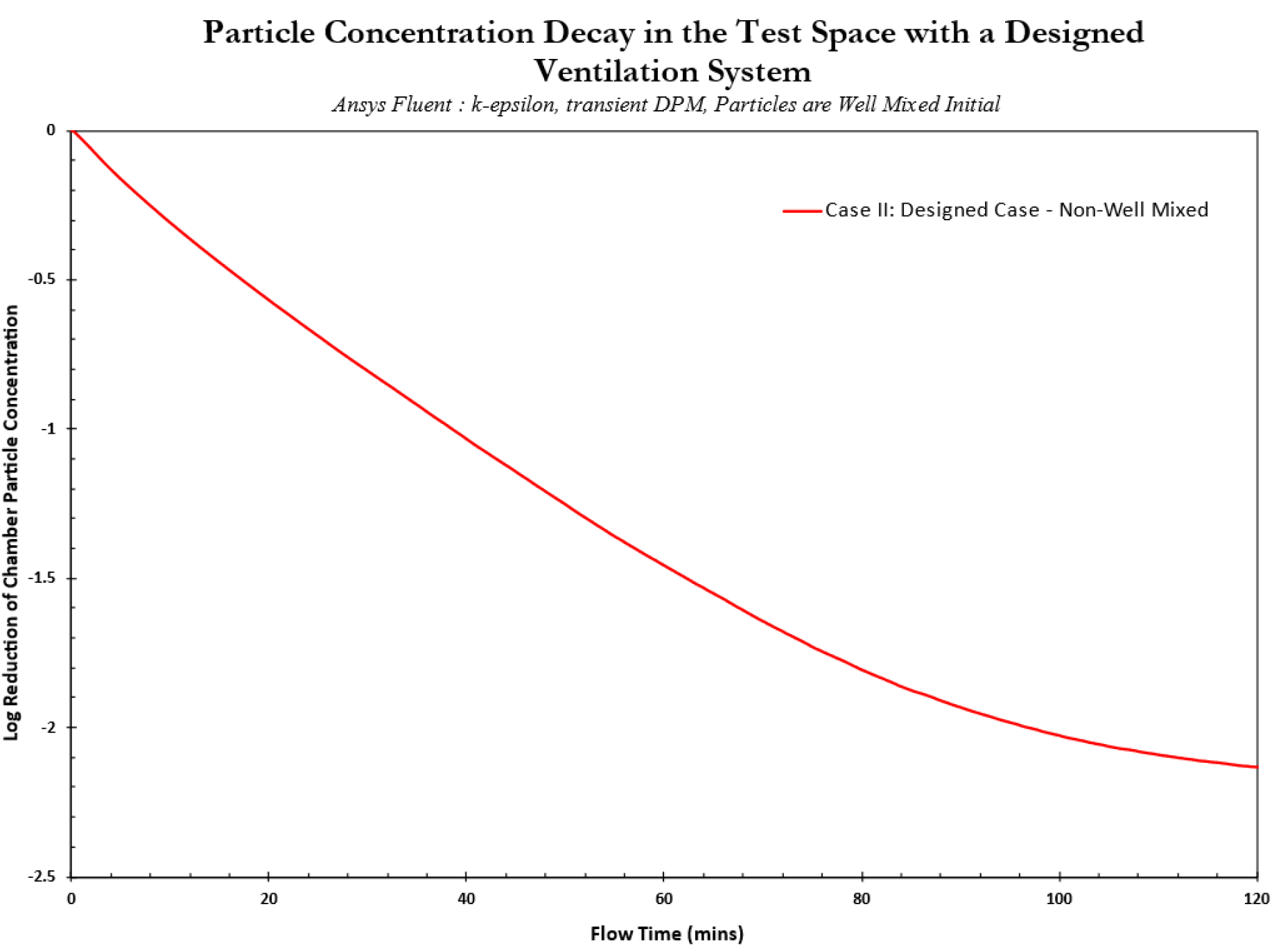
Total reduction of particles inside the designed ventilation system test chamber with one inlet pipe and one outlet pipe, with mixing fans placed inside the room. The airflow inside the room was designed to operate at 2000 CFM.

###### Brief Conclusion – Comparing the Standard HVAC and Designed Ventilation Case

By performing the above analysis, we observed that depending on the efficiency of the ventilation system (HVAC vs designed ventilation cases) and the degree of mixing in the room, there may be faster or slower clearance times that will vary depending on a variety of factors that are challenging to easily predict or explain ahead of time. For example, the slower clearance in the HVAC non-well mixed condition (vs HVAC well mixed) may be explained by the non-uniformity caused by the action of the mixing fans disrupting particle concentration uniformity across the volume. However, the faster clearance time seen for the well-mixed designed ventilation case (i.e. vs designed ventilation non-well mixed case) is producing this outcome through the homogeneity created by the mixing fans. Figure 15 compares the results between the designed HVAC case and the designed ventilation case for the test space. It is also clear that the standard HVAC system is also more efficient for aerosol removal under both mixed and non-well mixed conditions than the designed ventilation system, as would be expected.

**Figure 15.**
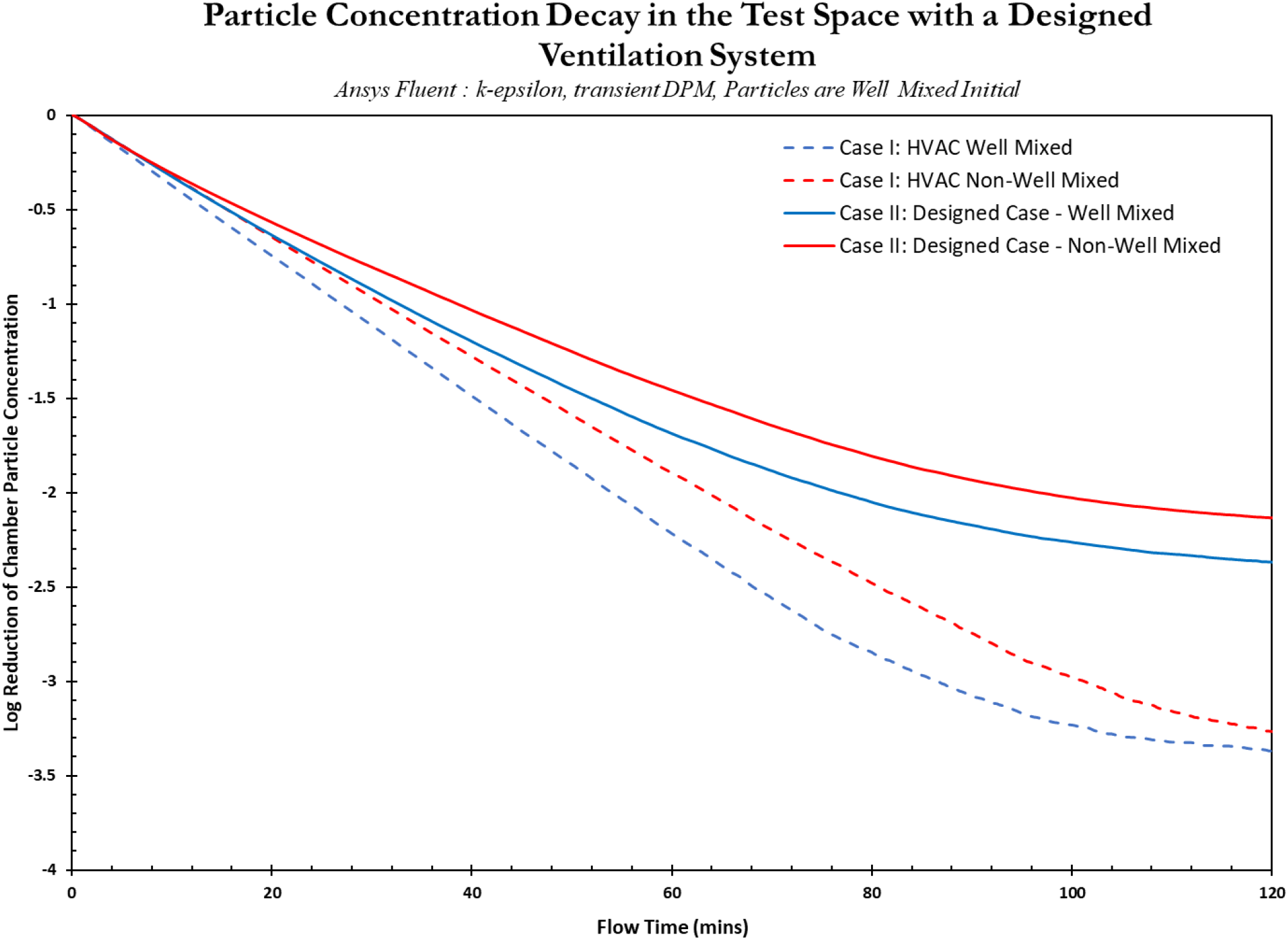
Comparison between the particle concentration decay for standard HVAC systems and the designed ventilation system for the test space under well mixed and non-well mixed conditions.

#### 3.3.3 Case III: The Realistic Case – Point Particle Releases

In the previous cases, we used uniform loading of particles across the volume of the virtual space to understand how results from a standard HVAC system design would compare to a designed ventilation system. In real scenarios, respiratory aerosol particles are released from a single source (i.e., an infected individual) and then they spread in the space following air paths – this process is best represented by the point release method in CFD. Based on the standards stated previously (ASHRAE and SMCNA), the flow rate inside the room was set to 2000 CFM. Again, the room was modeled to have one inlet pipe and one outlet pipe placed strategically within the test space. Twenty-three (23) virtual sensors were modeled into the room and placed in a zonal model around the point source (i.e. simulating a nebulizer) (Figure 16.).

**Figure 16.**
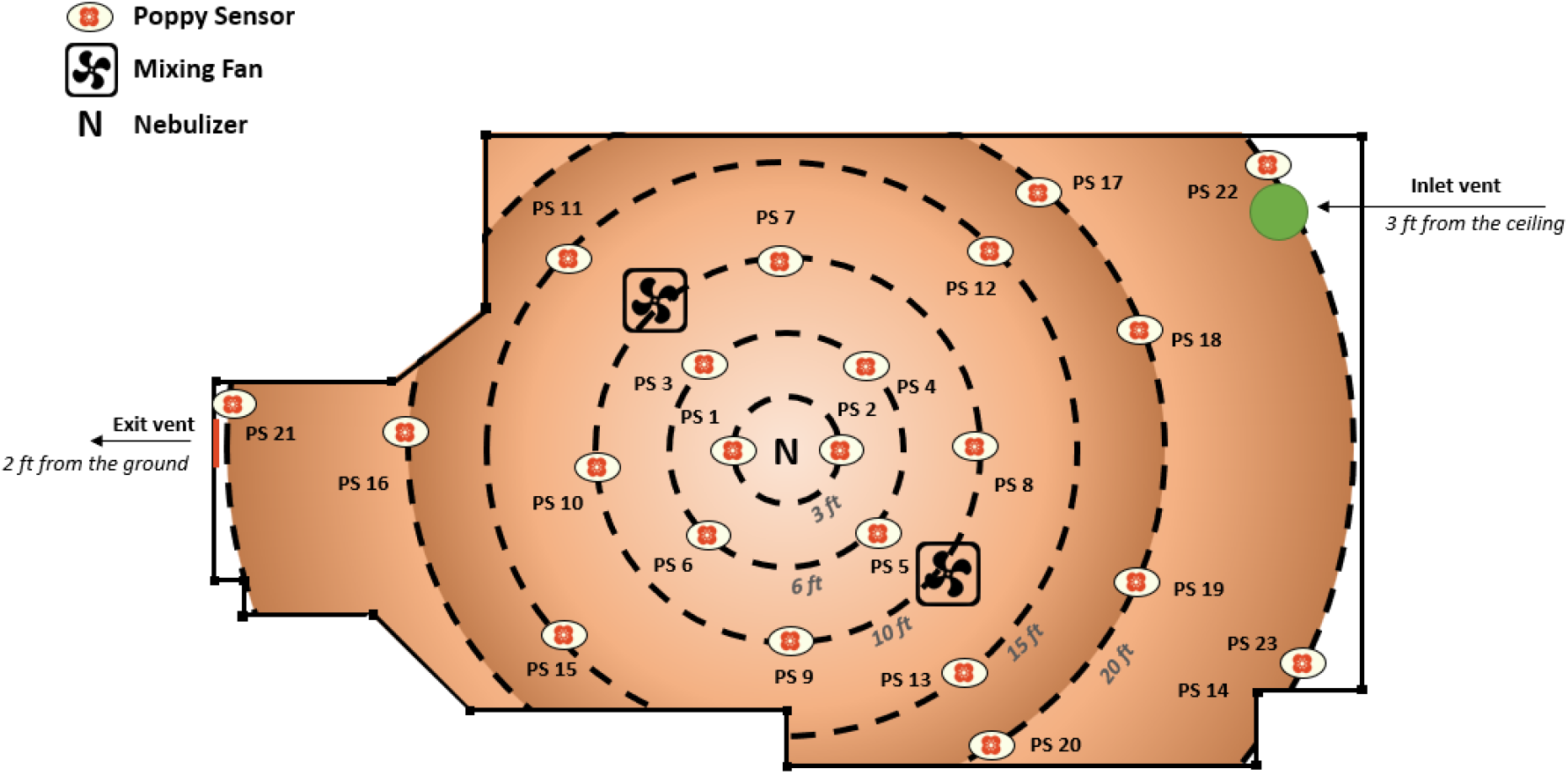
The geometric setup of the realistic ventilation case with one inlet pipe, one outlet pipe unit and two mixing fans placed inside the room. This is the extension to the case from the previous phase with 23 simulated Poppy Sensor devices to detect particles inside the chamber. The particles were modeled to release from the center of the room. The airflow inside the room was designed to operate at 2000 CFM.

As in previous cases, this study was performed to achieve predictions for two different flow conditions, well mixed and non-well mixed.

For the single-point release case in the CFD model, mixing fans are used strategically in both the well-mixed and non-well mixed conditions. As described below mixing fans are required to create uniform distribution of particles, or to create heterogeneity, depending on the nebulization phase of the experiment. The main difference between the well mixed and the non-well mixed conditions is that the well-mixed chamber was first filled with particles that were allowed to stabilize due to mixing fan activity, prior to the ventilation system being turned on. In contrast, in the non-well mixed condition the ventilation system was active throughout the whole experiment, including during nebulization, and the particles did not achieve stabilized concentrations.

Well mixed conditions were achieved by:

● The mixing fans were operational during the whole experiment (from nebulization to end of test).
● The particles released from the center of the room were allowed to fill the room first without exiting the chamber.
● The particles were allowed to spread out inside the chamber for an initial period of several minutes, until the concentration stabilized.
● Inlet and outlet vents were switched on once the sensors read stable particle concentrations.
● Data is logged continuously but the test data started once inlet and outlet vents were switched on.

Non-well mixed conditions were achieved by:

● The mixing fans were switched on after nebulization to increase heterogeneity
● The inlet and outlet ports were operational before nebulization, and the chamber had a defined flow field.
● The particles were released from the center of the room, and the data logging began immediately after the particle release started.

The numerical study was conducted in two steps.

● Obtaining the steady state solution for the flow inside the test chamber; the steady state solution implied that the flow inside the chamber had been solved until a constant flow pattern was obtained.
● Based on the steady state flow patterns, a particle tracking operation was performed where 1 million particles were introduced into the chamber, assuming that they would flow along the already established fluid flow inside the room and would not be collected on the chamber floor. The properties of the particles can be seen in Appendix A.

A solution for the existing flow field was used from the previous section, and only particle tracking analysis was performed for this case.

##### Non-Well Mixed Condition

###### Particle Tracking Analysis

After confirming the results, a transient Discrete Phase Model (DPM) was employed onto the steady state solution. The cone particle injection type was used to simulate the point particle release. A total of 1 million particles were required to perform this tracking exercise, and the release duration was set to 10 minutes. As ten different particle sizes were being released, the point release model was set to release 1667 particles of each size every second. The total simulation was performed for 120 computational minutes. The reduction in the concentration of the particles inside the chamber during the total simulation can be seen from Figure 17.

**Figure 17.**
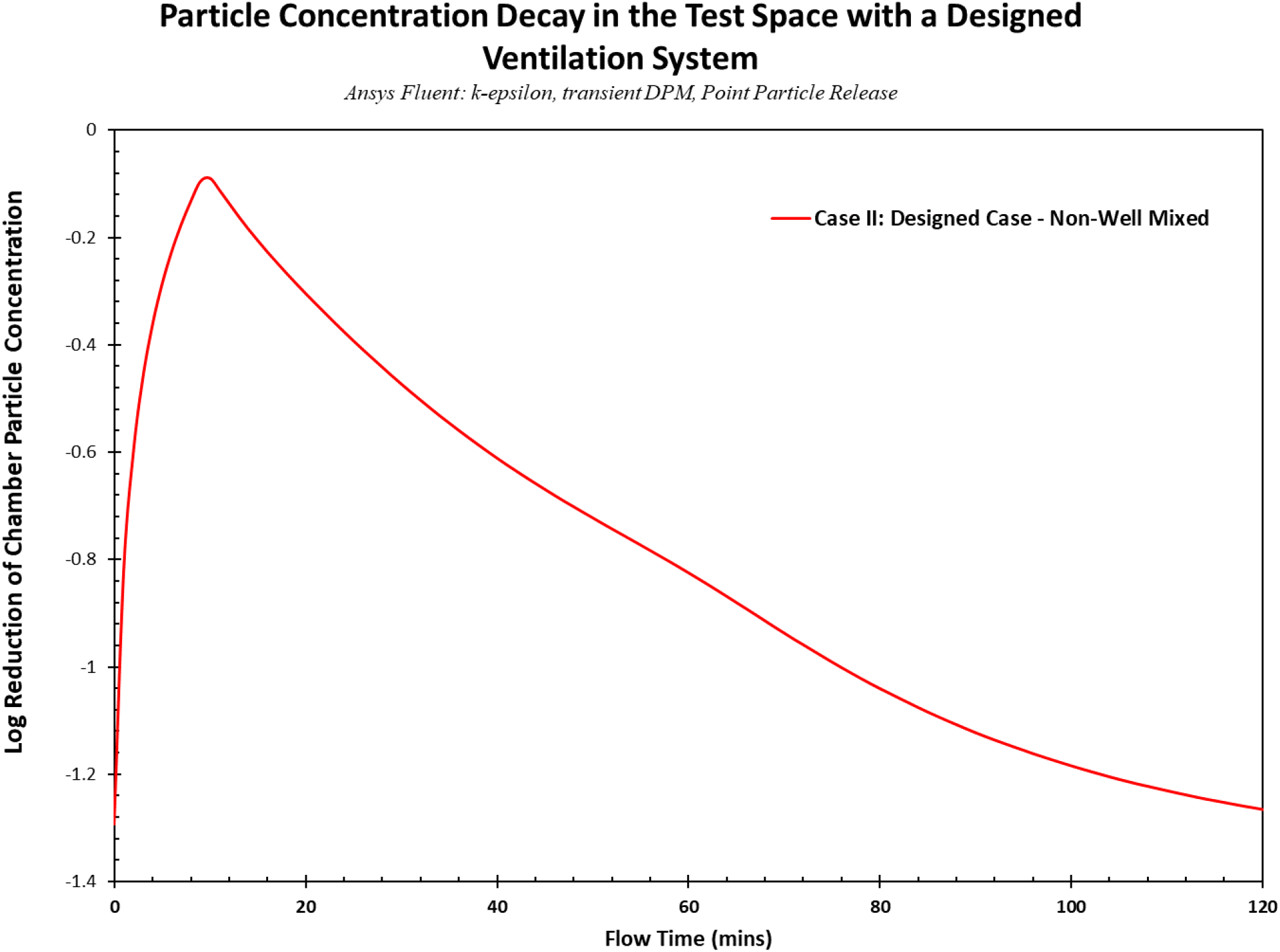
Total reduction of particles inside the test chamber with one inlet pipe and one outlet pipe unit placed inside the room. The particles were released from the center of the room, releasing 1 million particles in 10 minutes. The airflow inside the room was designed to operate at 2000 CFM.

It was observed that the particle concentration inside the chamber increased for the first 10 minutes and then started to go down as the particles moved along the flow of the air and started exiting the room through the outlet pipe of the chamber. The total reduction seen inside the chamber was 1.26 Log in 120 minutes (Figure 17).

##### Well Mixed Condition

###### Steady State Solution

A turbulent k-ε model was set up using ANSYS Fluent with the air coming out of the inlet pipe, whose diameter was 20 inches, with a flow velocity of 916.37 ft/min or 4.68 m/s. The walls were set to the no-slip boundary condition (which meant that the velocity at the walls was zero). The model had an outlet pipe with an outer diameter of 20 inches, which was set as a pressure outlet to balance the system. The flow inside the chamber was a constant 2000 CFM. Two mixing fans were introduced into the room (see Figure 18) to help the mixing inside the chamber. An internal fan boundary condition was used to simulate these fans’ operation, and they were set to operate at 750 CFM. The geometric setup of the modeled space can be seen in Figure 19.

**Figure 18.**
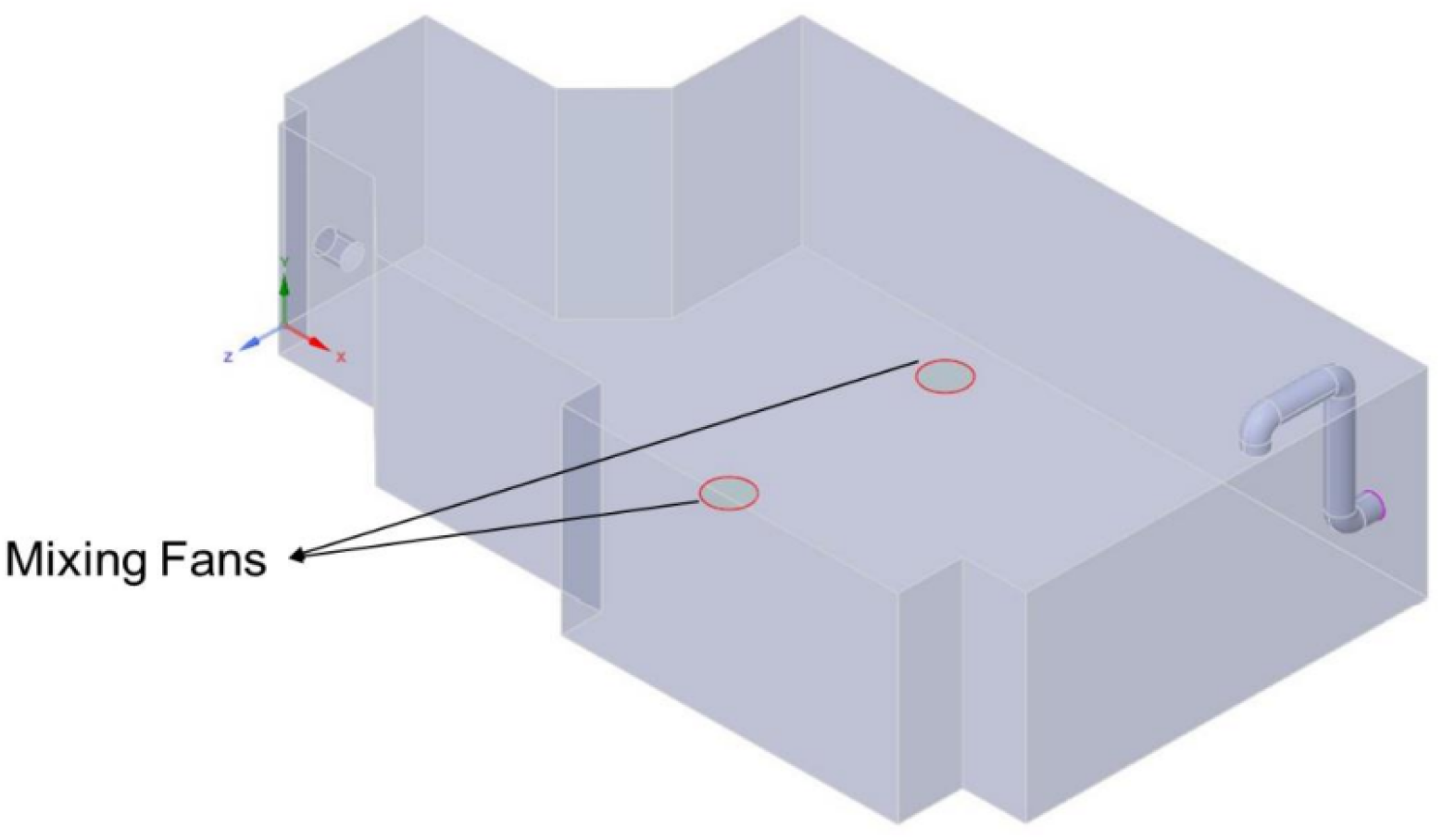
The geometric setup of the realistic case with one inlet pipe, one outlet pipe unit and two mixing fans placed inside the room. The airflow inside the room was designed to operate at 2000 CFM.

**Figure 19.**
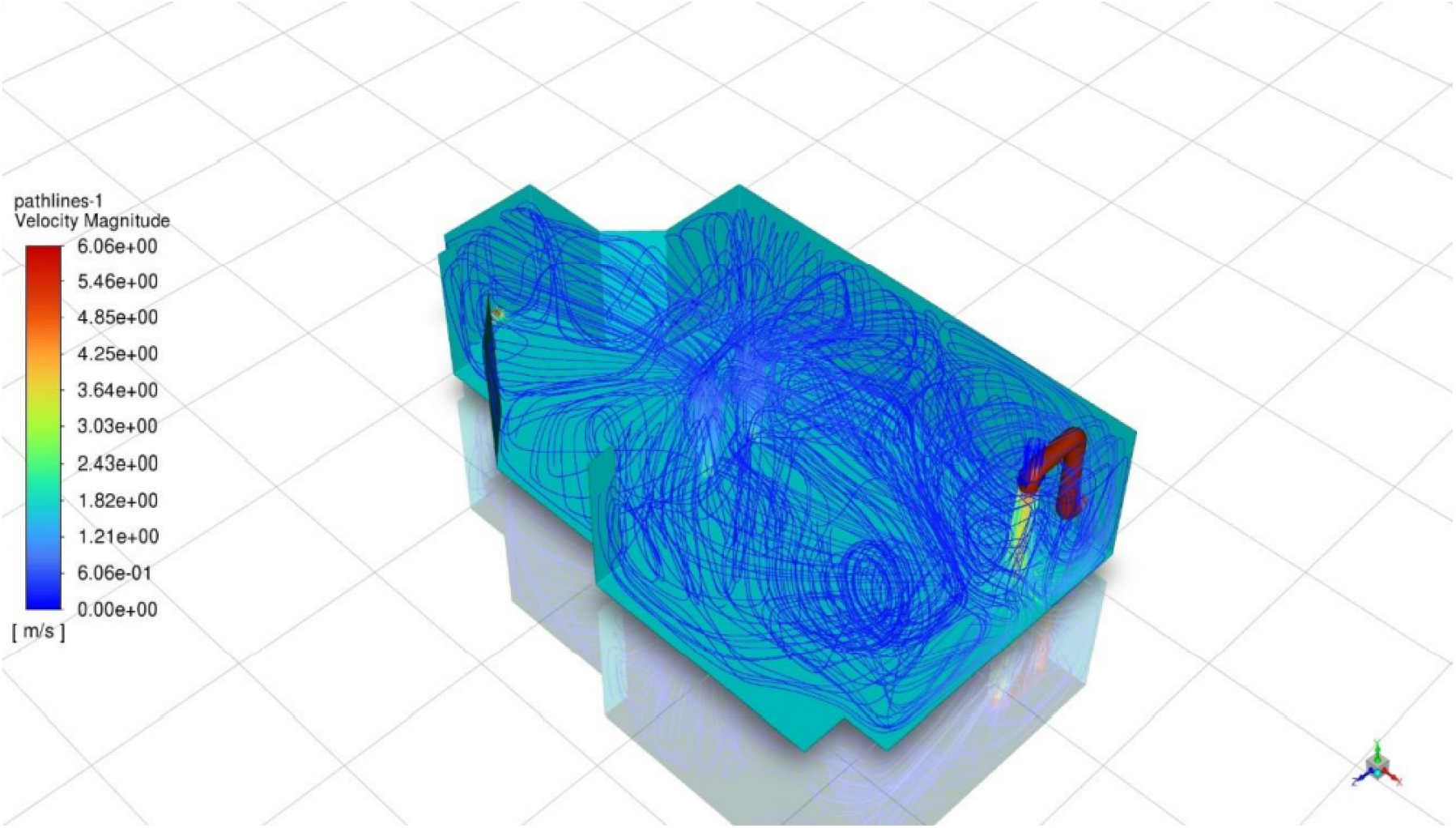
Flow inside the realistic case, numerically simulated chamber with one inlet pipe, one outlet pipe unit and with mixing fans placed inside the room. The airflow inside the room was designed to operate at 2000 CFM.

Watertight geometry workflow was used to perform the meshing of the CFD analysis. The standard local sizing and surface meshing settings were used to generate local, and surface meshes. Boundary layers were added at the walls of the geometry for accurate flow calculations at the walls, and a poly hexacore volumetric meshing was used to generate the computational cells. The meshing process generated an average of 1 million computational cells. The simulation was run until the solution satisfied a convergence criterion. The flow inside the chamber after getting a converged solution can be seen in Figure 20.

**Figure 20.**
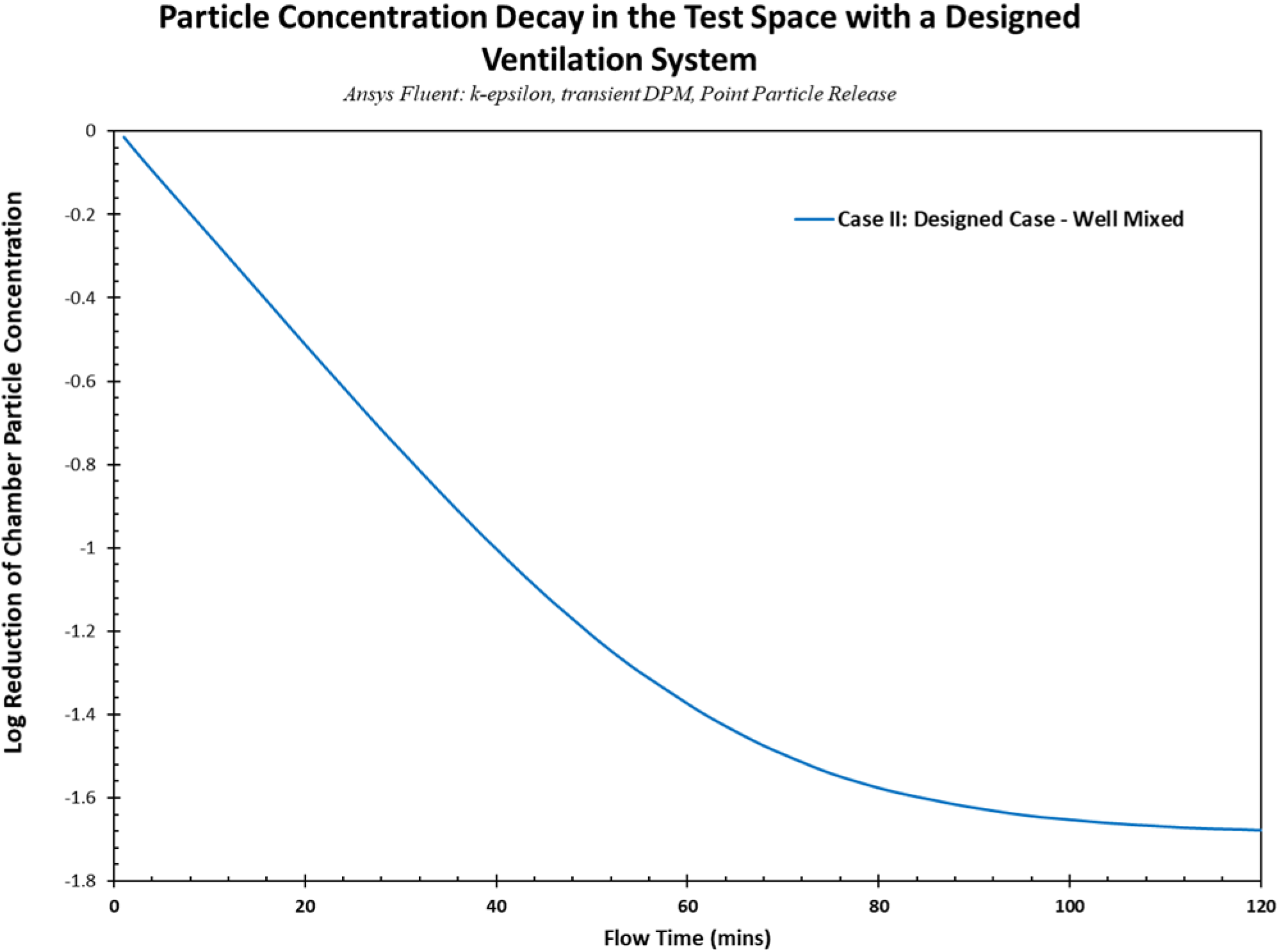
Total reduction of particles inside the realistic case test chamber with one inlet pipe, one outlet pipe and mixing fans placed inside the room. The particles were released from the center of the room, releasing 1 million particles in 10 minutes. The airflow inside the room was designed to operate at 2000 CFM.

Figure 19 shows the air entering the chamber from the inlet pipe, moving straight toward the floor. As it collides with the floor, it starts to diffuse into the chamber’s adjacent areas. Since the inlet duct was situated in the corner of the chamber, the diffused air collided with the walls adjacent to the flow to create local vortex zones. At the same time, the mixing fans are placed strategically inside the room to recirculate the local air around the fan to help the original flow from the inlet pipe to cover more area of the room. The flow eventually exits the chamber through the outlet pipe. From visual validation, the flow inside the chamber was as expected, and this solution was used as the set flow pattern for the particle tracking analysis.

###### Particle Tracking Analysis

After confirming the results, a transient Discrete Phase Model (DPM) was employed onto the steady state solution. The particles inside the chamber were introduced in the first-time step of the study, and the total simulation was performed for 120 computational minutes (Figure 20).

It was observed that the particles inside the chamber moved along the airflow and started exiting through the chamber’s return ducts. The natural particle reduction inside the chamber due to the flow field achieved a 1.68 Log reduction in 120 minutes, tested numerically (Figure 20).

###### Brief Conclusion

By performing the above analysis, we noticed that the mixed case had a slightly faster reduction inside the chamber than the non-well mixed case (Figure 22). This observation was in line with what was expected as the well mixed condition would mean that the particles would be spread out evenly and would exit the chamber faster than the non-well mixed case.

**Figure 22.**
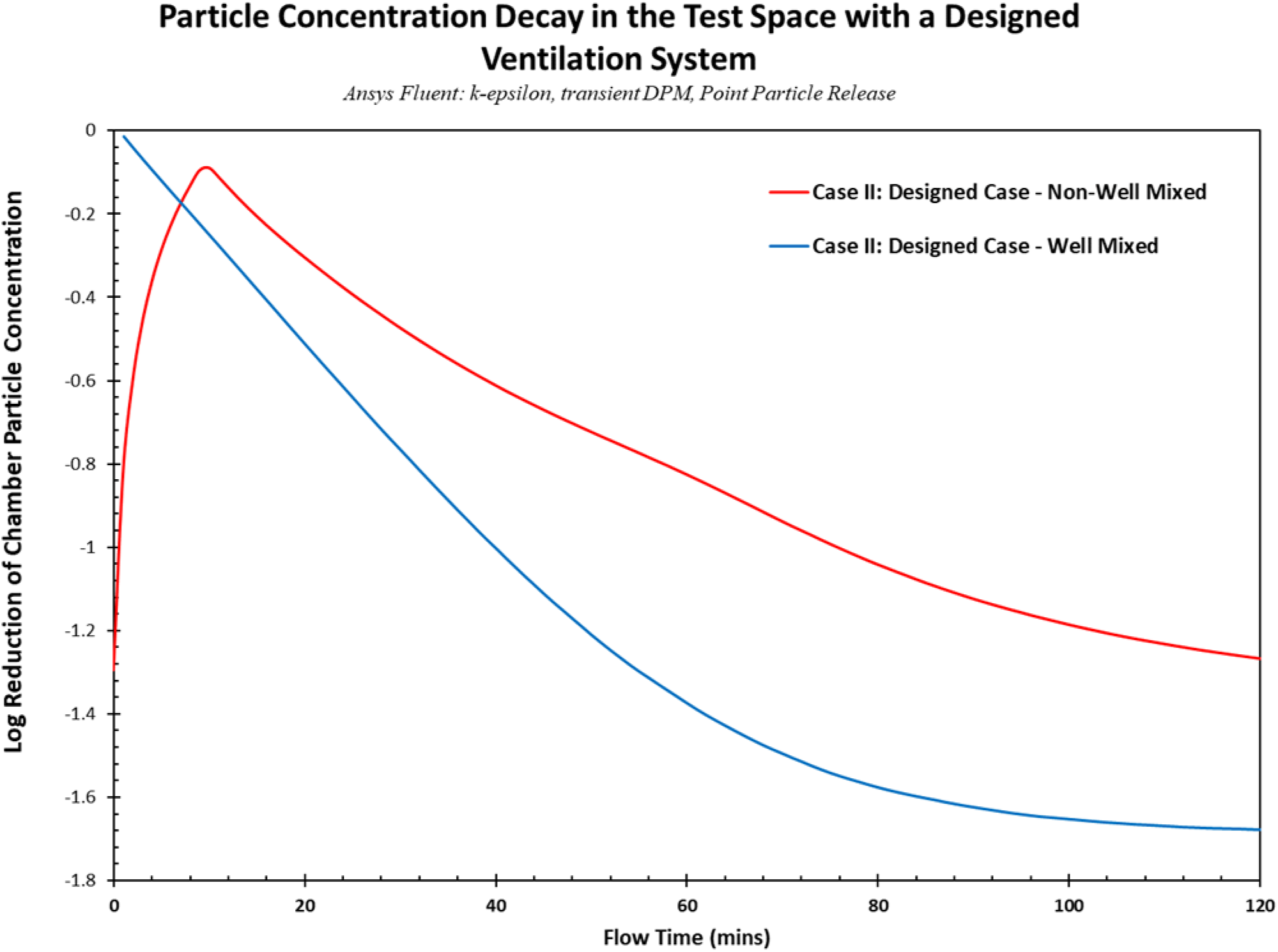
Total reduction of particles inside the realistic case test chamber with one inlet pipe and one outlet pipe, being operated to achieve a total room flow rate of 2000 CFM. The particles were released from the center of the room, releasing 1 million particles in 10 minutes, comparing the designed case with and without mixing fans.

##### Summary of the Analysis

This study simulated particle decay inside a large space with different HVAC system configurations using ANSYS Fluent. In case I, a large room was modeled using a standard HVAC system with ten supply and ten return channels, with a total airflow of 2000 CFM. The particles were volumetrically loaded, and the reduction inside the chamber due to airflow inside the room was analyzed. This case was studied in two configurations – without and with mixing fans. It was observed that the reduction inside the chamber was slower when the mixing fans were used as they increased the residence time of the particles inside the room. In case II, standard HVAC was replaced with a designed ventilation system that employed one inlet duct and one outlet duct operating at 2000 CFM. This case was also modeled without and with mixing fans. Comparing all the cases so far, we see that the standard HVAC case performed exceptionally well, achieving the highest reduction rate of 3.37 Log (Standard HVAC case without mixing fans) compared to 2.37 Log for the designed case (highest reduction rate achieved in the designed configuration – well mixed case). In our third study, the designed ventilation case was extended to have a point particle release from the center of the room (termed realistic case), and 23 Poppy Sensors were placed in a concentric zonal pattern to detect the particles at definite locations of the room. The mixing fans were also placed in the center of the room. It was seen that the total particle reduction inside the test chamber was higher during the well mixed case with 1.68 Log compared to the 1.27 Log achieved during the non-well mixed case. Overall, by performing these simulations, a baseline understanding of the flow field inside the room could be understood, and the particle decays inside the room can be analyzed in various configurations. In general, we see that well mixed conditions display faster aerosol particle clearance rates. It must be noted that the results seen in these simulations were performed with many assumptions to simplify the simulation, which motivated us to next use physical testing to examine the point release case in a real-world setting without the constraints of numerical modeling.

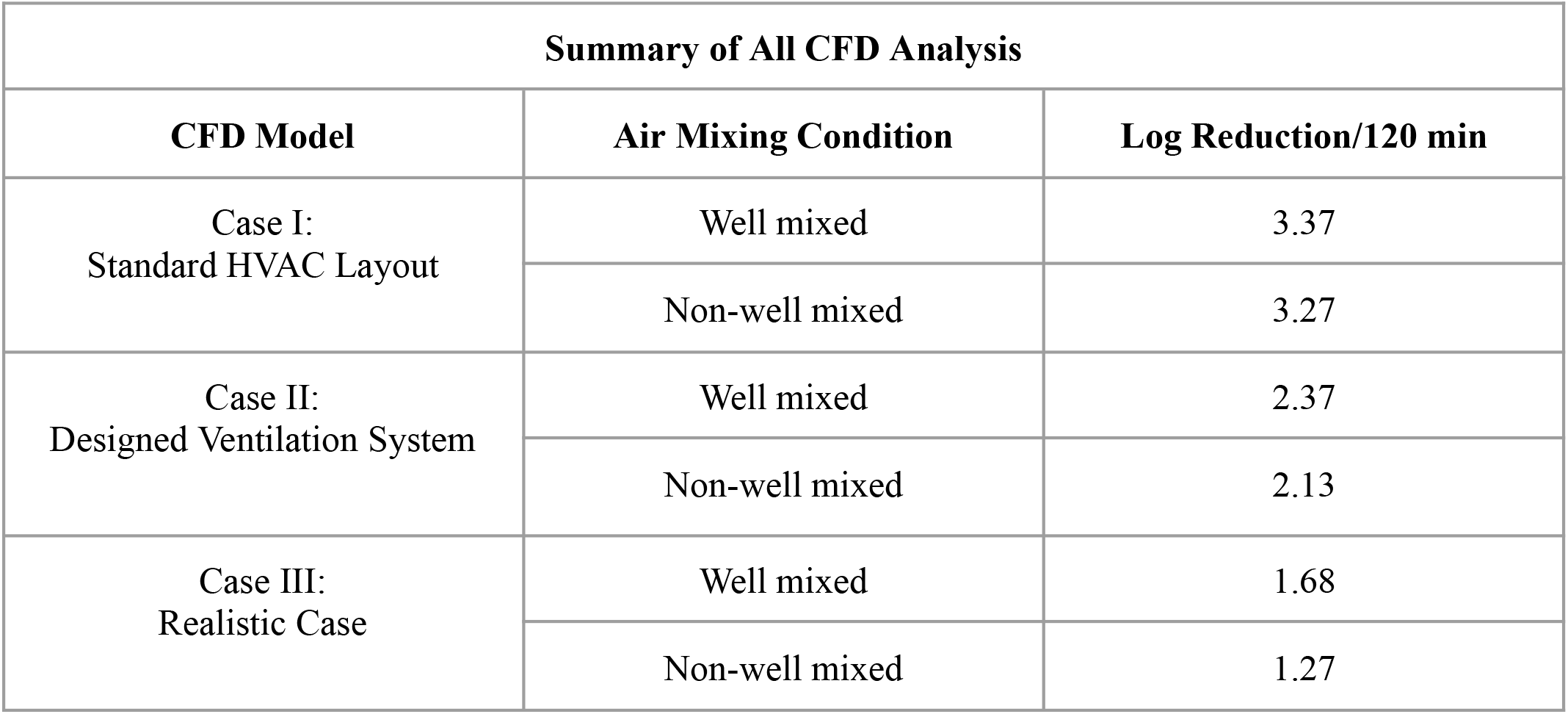

| Summary of All CFD Analysis |  |  |
| --- | --- | --- |
| CFD Model | Air Mixing Condition | Log Reduction/120 min |
| Case I:<br>Standard HVAC Layout | Well mixed | 3.37 |
|  | Non-well mixed | 3.27 |
| Case II:<br>Designed Ventilation System | Well mixed | 2.37 |
|  | Non-well mixed | 2.13 |
| Case III:<br>Realistic Case | Well mixed | 1.68 |
|  | Non-well mixed | 1.27 |

## 4. Physical Testing – eACH using Poppy Sensors in a Zonal Model

In the previous section, we used CFD modeling to simulate particle decay in a standard HVAC design and a designed ventilation system under well mixed and non-well mixed conditions. In the designed ventilation case we measured the local aerosol decay rate using 23 virtual sensors spread in concentric zones at a set distance from the nebulization source. While the numerical study enables us to compare ACH versus eACH measurements in a completely controlled system, the computational environment was not able to incorporate all physical processes that impact airflow and particle dynamics.

Thus, in the next section, we used a physical version of the designed ventilation system from our CFD study to examine these questions in more detail. Here, we utilized 16 Poppy Sensors spread out in a concentric zonal model, to monitor the concentration decay of tracer particles at each location.

### 4.1 Physical Test Setup

When defining minimum ventilation rates in buildings (except for low-rise residential), the key reference is [ANSI/ASHRAE Standard 62.1, Ventilation for Acceptable Indoor Air Quality](https://www.ashrae.org/technical-resources/bookstore/standards-62-1-62-2). This standard specifies minimum ventilation rates and other measures for both new and existing buildings intended to foster indoor air quality acceptable to occupants and mitigate adverse health effects.

Clean air only benefits occupants if it reaches them; for this reason, measurements of fine aerosol particle removal rates are most relevant within the “breathing zone”, and this is the area where the minimum ventilation rate is most important for occupant health. ASHRAE defines the breathing zone as the region within an occupied space between 3-72 inches above the floor and more than 2 feet from walls or fixed air-conditioning equipment.

A 24 jet Collison nebulizer with tracer liquid was placed at 3 ft vertically in the center of the test space. 17 Poppy Sensors were placed in concentric zones at a vertical height of 3 ft around the nebulizer, as shown in Figure 24. The first zone, situated at 3 ft from the nebulizer, had one Poppy Sensor placed opposite the nebulizer. The following zones (6 ft, 10 ft, and 15 ft from the nebulizer) had four Poppy Sensors, each set in the four cardinal directions with an offset of 45° between each zone. The final zone (20 ft from the nebulizer) had three sensors. The goal for this staggered pattern of device placement was to achieve maximum coverage of the area using the devices. As described previously, mixing fans in a physical test set-up are required to produce the well mixed condition of a more uniform distribution of air. Thus, two mixing fans were placed in the room to induce mixing to achieve a well mixed condition.

**Figure 24.**
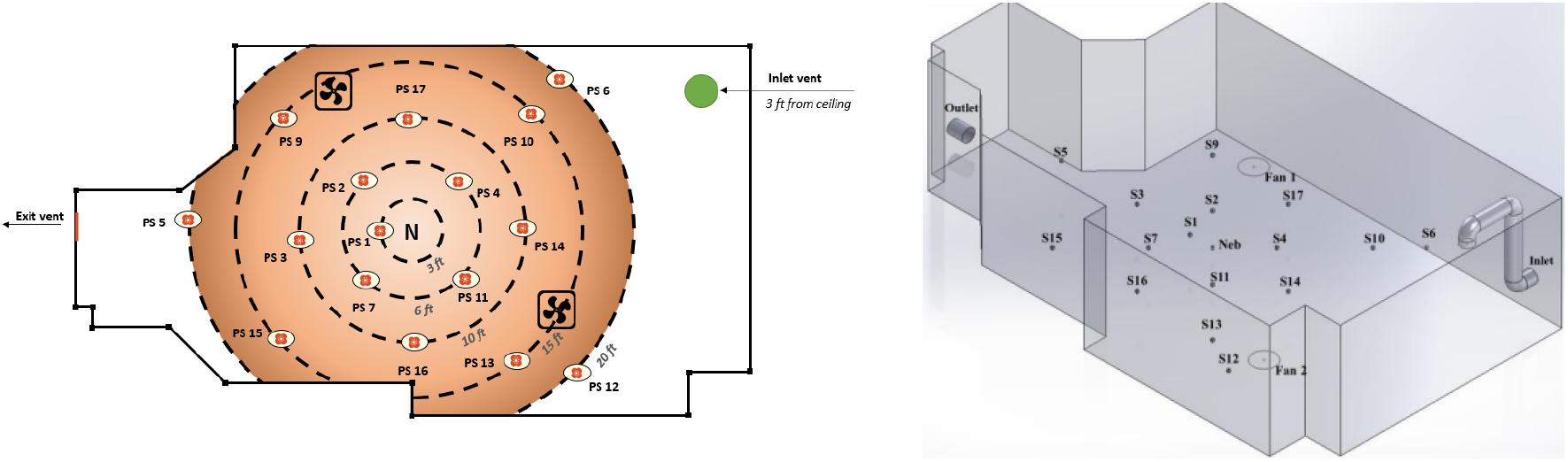
The test room setup. 16 Poppy Sensors (numbered 1-17 with #8 absent from the set due to data drop out for that device) deployed in a concentric zonal model around a diffuser with tracer liquid. The air was pushed inside the room from the end of the room using the carpet blower and flexible tubing. The air was pulled out from the opposite end of the room. The two mixing fans were placed in the room to achieve the well mixed condition of the room.

The entryways inside the testing space were blocked off using plastic tarp material to avoid air diffusion into unaccounted areas of the room. The inlet and outlet doors were sealed off with Masonite hardboard. The inlet and the outlet boards had 18” diameter holes so that the inlet duct could pass through the inlet board and the outlet blower could be attached to the outlet board. The inlet blower was placed outside the testing area to constantly provide fresh air into the testing chamber through a 25 ft long 18” diameter flexible duct (this was used for convenience over the 20” diameter duct that was specified in the CFD studies, due to material availability). The duct outlet was adjusted, so the air was released 3 ft below the chamber’s roof. The mixing fans were connected to a remote switch enabling hands-off operation. An air compressor was used to generate the pressurized air required for nebulization. The Poppy Sensors were connected wirelessly to a backend cloud system, where data was logged to the cloud every 15 seconds.

### 4.2 Materials/Equipment

The Poppy Sensors (Figure 25) used in this study provide real-time tracer particle concentration measurements by connecting to a wireless access point remotely. These devices have a particle count accuracy of ± 10%, with a resolution of 1 tracer particle per liter of air. The counted tracer particles range from 0.1µm to 10µm in particle size, with differential binning for tracer particle counts, allowing the 1μ*m* particle count fraction to be isolated and analyzed during these experiments. This is important because human respiratory aerosol emissions often present a peak in their size distribution at close to 1 μ*m*. Therefore, when calculating the eACH for a space we are interested in the effective removal rate for this representative fine aerosol particle fraction. The air sampling velocity of the sensors is 0.125 LPM.

**Figure 25.**
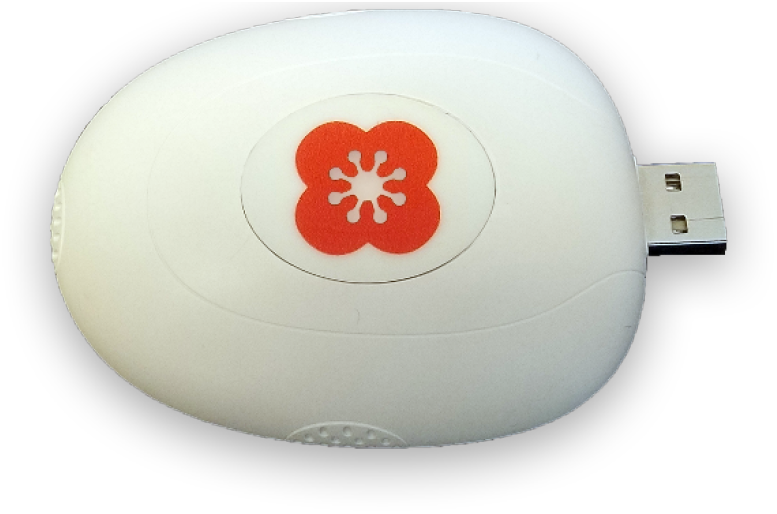
The Poppy Sensors used in this study. A total of 16 sensors were employed in the test space in a concentric zonal model around a diffuser with tracer solution.

Tracer particles were disseminated using a Collison 24-jet nebulizer (BGI Inc. Waltham MA), similar to the model shown in Figure 26. The aerosolization of the tracer solution was driven by a filtered air supply generated by a pump. A pressure regulator allowed for control of disseminated particle size, use rate, and sheer force generated within the Collison nebulizer. Before testing, the Collison nebulizer flow and use rates were characterized using an air supply pressure of approximately 40-60 psi. This produced an output volumetric flow rate of 50-80 L/min with a fluid dissemination rate of about 1.25 mL/min. The Collison nebulizer was flow characterized using a calibrated TSI model 4040 mass flow meter (TSI Inc., St Paul, MN).

**Figure 26.**
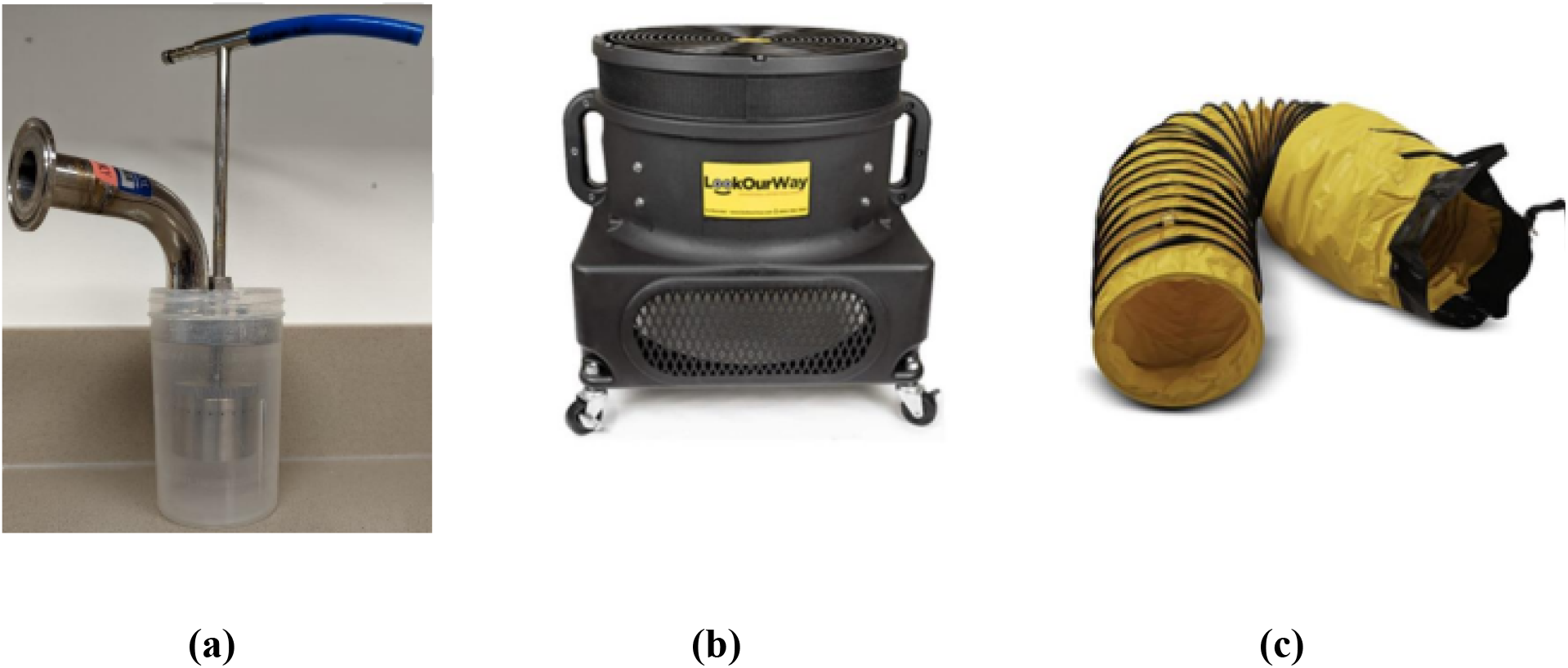
Materials used in test space: **(a)** 24-jet Collison nebulizer with 304 stainless steel construction (BGI Industries). **(b)** The 1HP blower used to move air within the test chamber. **(c)** The 18” diameter flexible duct used to bring in fresh air through the inlet.

An 18” blower (Figure 26-b) was used as an inlet and outlet for testing. The blower came equipped with a 1HP AC fan with three different fan settings. The other speed settings planned for this study were achieved by blocking the intake grills. An 18-inch flexible tube (Figure 26-c) was used to direct the supply from the inlet into the testing chamber.

### 4.3 Test Matrix

To provide airborne infection control, selecting the appropriate target air change rate is one of the most crucial decisions for a facility. Too few air changes per hour increases exposure levels and infection risk for occupants. Too many, on the other hand, unnecessarily increases facility costs from purifiers, energy usage, and mechanical strain, thus creating additional economic strain and an increased carbon footprint for the building.

The CDC recommends ACH levels to help clinical facilities maintain effective infection control practices. Recommended ACH values range from 2 to 12 or more, depending on the function of a particular space. The American Industrial Hygiene Association (AIHA) similarly recommends using engineering controls in all indoor workplaces, including those outside the healthcare industry, to reduce the spread of COVID-19. The ACH values in the non-healthcare sector vary between 2-15 ACH according to the size, occupancy, and intended activities of a space. Based on this, we tested five air change rates, within the range of 0 – 8.7 ACH. This range was selected to span the LANCET COVID-19 Commission’s <u>proposed non-infectious air delivery rates</u> of 4 (good), 6 (better), >6 (best) ACHe for typical non-residential spaces. **Table 1** (below) summarizes the test cases performed as a part of this study.

**Table 1.** Testing five (5) different flow rate configurations. For each set flow rate, testing was performed for two different conditions: non-well mixed and well mixed conditions. The 2060 fpm case (8.70 ACH) was performed six times (three for non-well mixed and three for well mixed) to test the replicability of the tests.

| Room Flow Configuration | Flow Rate Setting (Calculated ACH) |  |  |  |  |
| --- | --- | --- | --- | --- | --- |
|  | 0 FPM<br>(0 ACH) | 520 FPM<br>(2.20 ACH) | 1020 FPM<br>(4.31 ACH) | 1470 FPM<br>(6.21 ACH) | 2060 FPM*<br>(8.70 ACH) |
| Well Mixed | ✓ | ✓ | ✓ | ✓ | ✓ |
| Non-Well Mixed | ✓ | ✓ | ✓ | ✓ | ✓ |

### 4.4 Experimental Procedure

Mechanically supplied ventilation can supplement low natural ventilation to a room, to ensure abundant air exchange and mixing for infection control. While it is generally accepted that mixing is critical for well-ventilated space, the addition of occupants and items in the space tends to create localized zones and cause a heterogenous flow system. Two conditions were tested experimentally at each flow rate setting to understand the effect of uniform and heterogenous mixing on effective air change rates.

#### (a) Well Mixed Condition

The goal of conventional fully mixed air distribution is to create a “well mixed” environment which essentially means that the concentration of particles released at a point will tend to spread uniformly within the room. To achieve this, mixing fans were employed to propel the particle from the source to the different corners of the room. Following is the experimental procedure for the well mixed condition:

1. The Poppy Sensors record background tracer particle concentrations for 5 minutes.
2. The mixing fans were switched ON, and the blowers (ventilation) were switched off.
3. The nebulizer with tracer liquid was switched on for 10 minutes.
4. The nebulizer was turned OFF after 10 minutes, and the nebulized particles were allowed to travel across the volume of the room naturally or induced by airflow from the mixing fans for the next 10 minutes. This was confirmed by observing stable particle counts on the Poppy Sensors for 1 minute.
5. The blowers were turned ON and set to run until the particle counts reached the background levels observed before starting the next trial.
6. The trials were repeated with different flow speeds.

#### (b) Non-Well Mixed Condition

Heterogeneous air distribution patterns affect the uniformity of an indoor particle concentration field. To create a non-uniform distribution of particles, the system is not allowed to saturate, and particles are nebulized in a chaotic airflow induced by the mechanical ventilation in the designed case. Note that no mixing fans are used in this case. The following is the experimental procedure for a non-well mixed condition:

1. The Poppy Sensors record the background tracer particle concentrations for 5 minutes.
2. The blowers (inlet and outlet) were switched on, allowing the air to flow through the test space.
3. The nebulizer with tracer liquid was switched on for 10 minutes.
4. The nebulizer was turned OFF after 10 minutes.
5. The Poppy Sensors sampled the particles. The study was considered finished when the Poppy particle detectors reached the background value recorded at the start of the test
6. The trials were repeated with different flow speeds.

### 4.5 Data Analysis

In a study by Johnson et al. (2011), it was reported that healthy subjects (8–15 humans) generate respiratory aerosols over three size modes during speech and voluntary coughing (1.6, 2.5 and 145 μm, and 1.6, 1.7 and 123 μm, respectively) [11]. These particles contained very large respiratory droplets with sizes exceeding 100 μm, which fell to the ground within a few seconds. However, in experimental trails, small particles of approximately 1-2 μm were generated simultaneously [11], and they could remain airborne for long periods of time. The Poppy Sensors collected the raw particle counts and concentration of nebulized tracer particles of sizes ranging from 0.1 µm to 100 µm. For this study, only the 1 µm particle size was considered for the analysis because it is close to the 1.6 um size mode for respiratory aerosols. The data was collected in real-time and then processed computationally. Under the assumption of well mixed conditions and no sources of emission, the mass balance for a substance in space is described as follows:

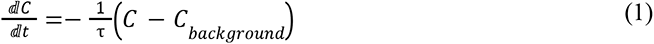

Here *C* is the concentration of the tracer substance in the room, *t* is time, τ is the air change timescale, and *C _background_* is the background concentration of the tracer substance in the room when ventilation is operated at a specified condition, but no tracer substance has been released. By introducing a corrected concentration variable, *Ĉ*= *C − C _background_*, Eq. (1) simplifies into:

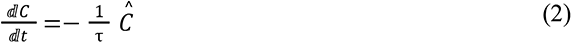

Thus, τ can be determined as the inverse slope of the linear regression line of the natural logarithm of *Ĉ* Versus time. Note that if sources of the tracer compound are present, the *C _background_* may vary with time, resulting in the nonlinearity of this relationship. Once exponential decay is confirmed, analysis can be simplified by calculating τ as the *e*-folding time or the time for the peak concentration to be reduced by a factor of *e*. We define eACH as follows:

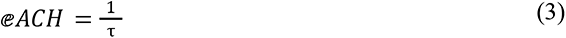

## 5. Results

### 5.1 Numerical study – eACH from virtual sensors

To mimic a realistic condition, a point release numerical study was conducted in the test space with the designed ventilation system. Based on the standards stated previously (ASHRAE and SMCNA), the flow rate inside the room was set to 2000 CFM (4.78 ACH). The room was modeled to have one inlet pipe and one outlet pipe placed within the test space (Figure 9.). The room was set up in this way to achieve a realistic arrangement of components so that these simulations could be replicated in physical testing. Twenty-three (23) sensors were modeled into the room and placed in a zonal model around the source (the nebulizer). Particles were uniformly loaded at the start of the test and the numerical solver calculated the decay of the particles at these sensor locations.

Figure 27 shows the eACH captured by each of the 23 sensors at positions throughout the virtual test space. When these eACH values were compared to the positional air paths for the non-well mixed condition it is clear that eACH values deviated from the well mixed condition and average for the space according to local increases in air velocity. This is most evident for the highest velocity regions around the inlet and outlet vents (sensor 22 and 21, respectively).

**Figure 27.**
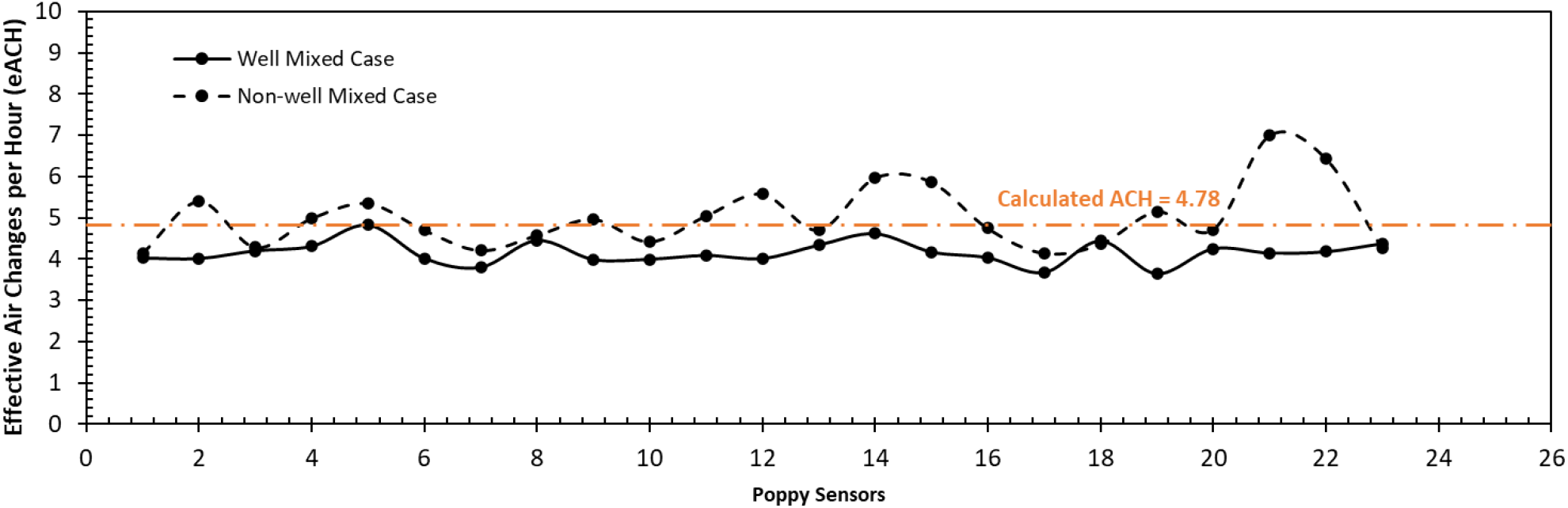
eACH captured by 23 sensor locations modeled in the virtual test space with a designed ventilation system for a well mixed and a non-well mixed case. The well mixed case had lower spatial variability than the non-well mixed case.

**Figure 28.**
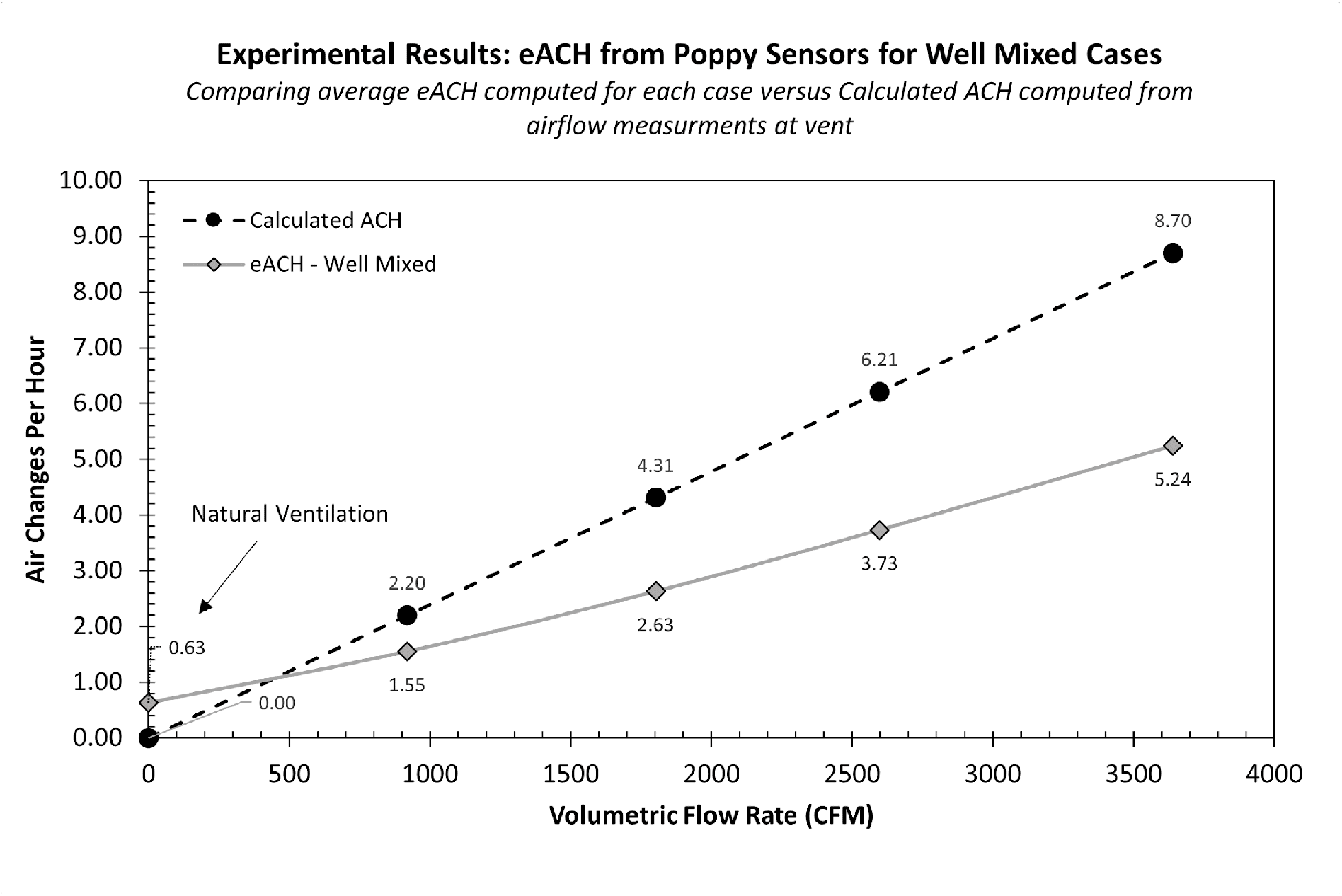
Calculated air change rate and measured effective air change rate versus the volumetric flow rate for well mixed cases in the physical test space. As increasing levels of air are exchanged through the test space, the effective ACH increases but at a slower rate. A deviation as high as 40% is noted.

**Figure 29.**
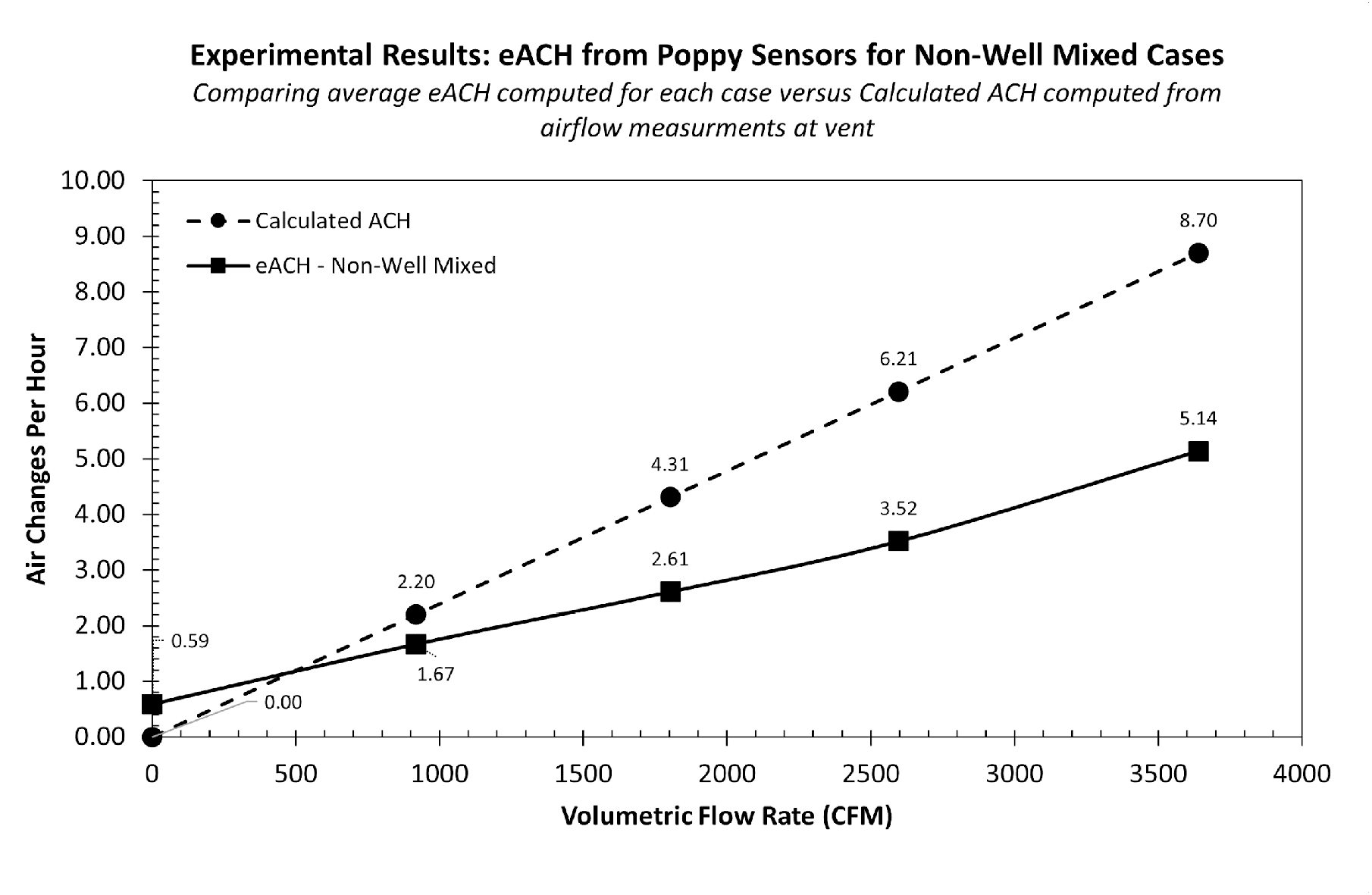
Calculated air change rate and average measured effective air change rates versus the volumetric flow rate for non-well mixed cases in the physical test space. As increasing rates of air are exchanged through the physical test space, the effective ACH increases but at a slower rate. A deviation as high as 43% is noted.

Given that the majority of real-world rooms are not well mixed, these results highlight the need for direct measurements of respiratory aerosol removal rates within the breathing zone of rooms at representative positions that are typically occupied during group use of a space. Further, when the positional eACH values for the non-well mixed condition were compared to the 4.78 ACH, which is the calculated ACH for the virtual space, the potential for meaningful local deviation in actual particle removal rates become apparent. Considering the difference in reduction of COVID-19 transmission risk at 4 ACH versus 6 ACH (66.8% versus 82.5% reduction, respectively) that was seen in the 2022 Italian Schools study [9], a loss of 1-2 effective air changes for respiratory aerosol removal could create significant local airborne infection risks in rooms with ventilation that has been been designed to reach a minimum of 4 ACH (for example) based on calculations alone.

### 5.2 Physical Study – eACH from Poppy Sensors

The physical test space was described in section 3.3. To calculate air changes per hour for this space, we measured: (a) Room volume – a laser distance measure (Bosch GLM165-40) was used to estimate the lateral and vertical dimensions and using those dimensions, the effective volume of the space was calculated as 25095 ft^3^; and (b) Volume of air out of the inlet – a rotating vane anemometer (AP856A, AOPUTTRIVER) was used to calculate the volumetric flow rate being supplied out of the inlet. Multiple measurements were taken at the inlet, and an average stable reading was calculated. During pre-testing the blowers were turned on, and settings were pre-marked on the blower controls to deliver a range of flow rates (520-2060 FPM). During testing, the flow rates were rechecked for accuracy. With these two measurements, the calculated Air changes per hour (ACH) were be computed as:

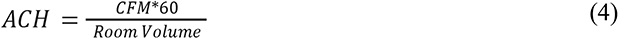

To understand the general mechanics of the particles in the test space, tracer particles were first nebulized in the test space without active ventilation. The room was undisturbed for a prolonged period to accurately represent zero ACH. For the non-well mixed case, tracer particles were nebulized without mixing fan activity, and the sensors recorded during the ramp-up and the decay period. For the well mixed case, the mixing fans were run during nebulization to induce mixing, followed by a 10 min settling period to enable the concentration to stabilize across the room uniformly as possible. Under the condition of zero ACH, an average eACH of *0.59* and *0.63* was calculated using 16 sensors deployed for the well mixed and non-well mixed cases, respectively. These numbers correspond to ACH levels often observed in homes. Brownian motion is why values between 0 -1 are generally seen under conditions of zero ACH.

The designed ventilation was next engaged to produce four increasing air change rates, while the 16 Poppy Sensors around the nebulizer source measured the decay of tracer particles from positions throughout the space. For the well mixed case, as we increased the volumetric airflow into the space, the rate of decay and the corresponding measured value (eACH) increased. It was also observed that the measured eACH values were lower than the theoretical calculated ACH value, and this deviation increased as we increased the flow speed. On average, except for the 2.20 ACH case (29.54 % deviation), the theoretical and derived values differed by 39.60 % (Table 2).

**Table 2.** Calculated ACH and measured Effective Air Change Rates for well mixed cases in the physical test space.

|  |  |  |  |  |  |
| --- | --- | --- | --- | --- | --- |
| Measured Value (FPM) | 0 | 520 | 1020 | 1470 | 2060 |
| Flow rate (CFM) | 0 | 918.91 | 1802.49 | 2597.70 | 3640.32 |
| Calculated ACH (hr<sup>-1</sup>) | 0 | 2.20 | 4.31 | 6.21 | 8.70 |
| Measured eACH (hr<sup>-1</sup>) | 0.63 | 1.55 | 2.63 | 3.73 | 5.24 |
| Difference (%) | - | 29.54 | 38.97 | 39.93 | 39.77 |

Similarly, for the non-well mixed case, the rate of decay and the corresponding derived value (eACH) increased as we increased the volumetric airflow into the space. On average, except for the 2.20 ACH case (24.09 % deviation), the calculated ACH vs experimentally measured eACH values differed by 41.23 % (Table 3).

**Table 3.** Calculated air change rate and average measured effective air change rates for non-well mixed cases in the physical test space.

|  |  |  |  |  |  |
| --- | --- | --- | --- | --- | --- |
| Measured Value (FPM) | 0 | 520 | 1020 | 1470 | 2060 |
| Flow rate (CFM) | 0 | 918.91 | 1802.49 | 2597.70 | 3640.32 |
| Calculated ACH (hr<sup>-1</sup>) | 0 | 2.20 | 4.31 | 6.21 | 8.70 |
| Average Measured eACH (hr<sup>-1</sup>) | 0.59 | 1.67 | 2.61 | 3.52 | 5.14 |
| Difference (%) | - | 24.09 | 39.44 | 43.31 | 40.91 |

Both well mixed and non-well mixed for each flow rate showed a significant deviation (∼40% for >4 ACH, 24% for 2.20 ACH) from the calculated ACH for the room. The calculated ACH value derived by measuring the volumetric flow rate and a standard formula is part of general practice by HVAC professionals. While this is a common practice (ASHRAE 41.2P), this type of measurement is often unreliable, and is dependent on several factors with high levels of variation from measurement to measurement, such as the direction and precise location of the measurement. The development of static pressure in the vents can also lead to unstable measurements. Depending on the flow regime (fully developed or underdeveloped), the flow can be laminar or turbulent affecting the measurement. This may be one of the reasons that 2.20 ACH showed a lower deviation in both the well mixed and non-well mixed cases. Previous studies [16, 17] have highlighted the inaccuracy found in the field measurements of airflow at HVAC grills. For this study, multiple stable measurements were taken, and an average was considered as the “Measured Value (FPM)” for our calculations.

Xu (2014) highlighted the non-uniformity caused by the placement of inlet(s) and the type of inlet [18]. In their study, Cui et al. (2015) used CO_2_ as a tracer gas to measure the air change rate in a highly controlled 636 ft^3^ chamber [19]. A maximum error of about 15% was estimated in calculating the ACR at the inlet. The flow was fully developed in the vents before releasing in the experiment, and an average deviation of 12 % was recorded. They also noted that spatial variation was present, indicating that imperfect mixing is unavoidable. Westgate and Ng (2022) expanded on this study by using in-situ CO_2_ and PM measurements to assess air change rates [20]. They found similar trends to the latter but noted that PM decay measurement also varied with the deposition of the particles. Figure 30 compares the eACH in well mixed and non-well mixed cases. Both cases yielded similar results, highlighting the conclusion of imperfect mixing found by Cui et al. (2015).

**Figure 30.**
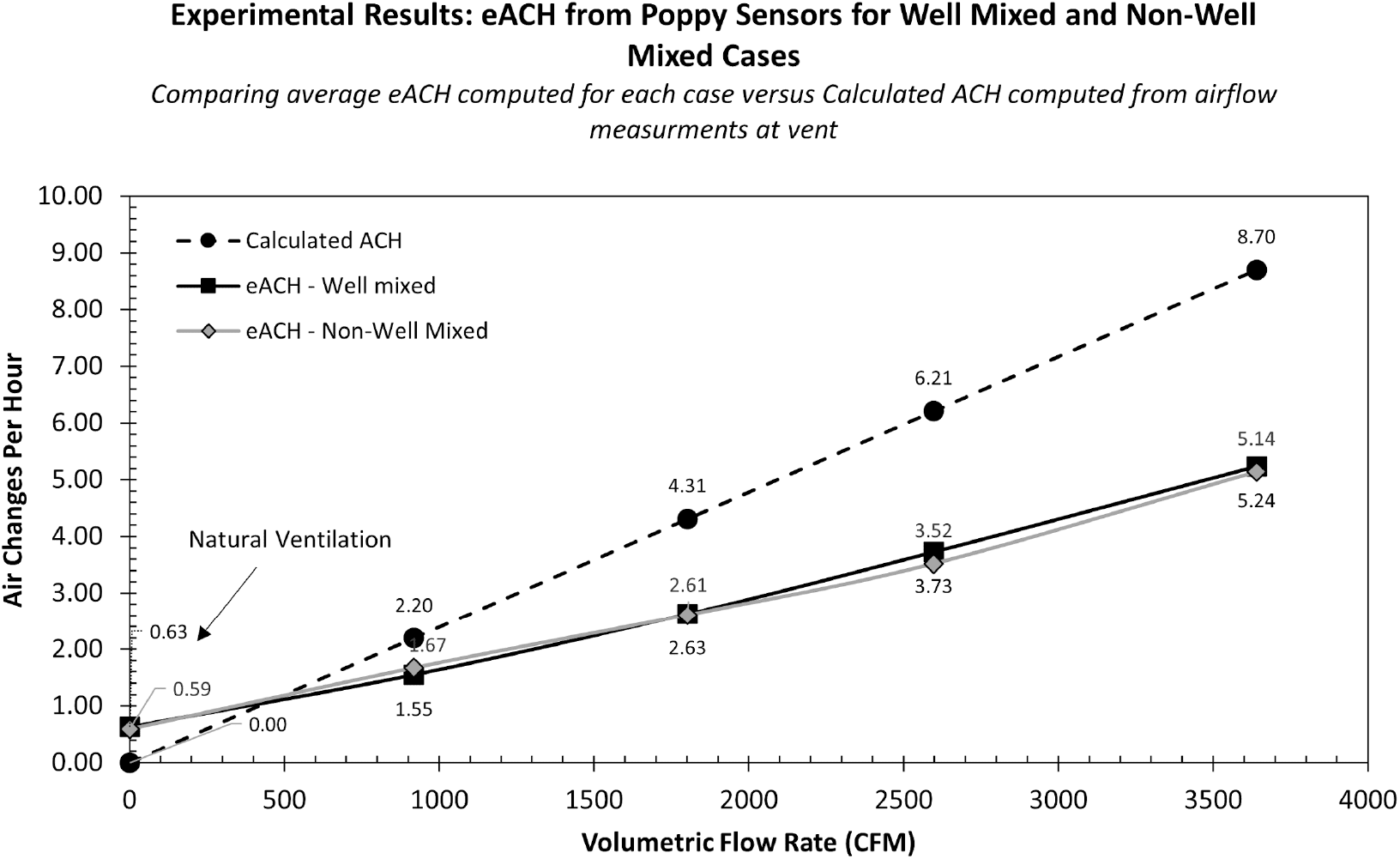
Calculated ACH & measured eACH versus the volumetric flow rate for well mixed and non-well mixed cases in the physical test space.

Figure 24 shows the experimental setup where 16 Poppy Sensors spread out in a concentric zonal model, mapping the concentration decay of particles at each location. Different ventilation systems and designs lead to variable airflow patterns in a room. While a fully uniform system is desired, zones that are highly ventilated and under-ventilated are common. Spatial variation can be caused by localized recirculation zones, which leads to variability in tracer concentrations and decay associated with it over time. Figure 31 shows how the value of eACH varies spatially around the source (nebulizer) for five different volumetric flow rates in the test space. Detectors closer to the inlet, outlet and walls tend to have higher eACHs as particles in those zones both have higher velocities while also experiencing the effects of greater surface area near the walls for particle deposition. As the volumetric flow rate increases the spatial eACH variability also increases, likely due to elevated turbulent flow in the room, creating pockets where faster or slower decay occurs. These results recapitulate the spatial variability effects that were observed with the numerical study, albeit with less separation between the well mixed and non-well mixed conditions. The spatial variability found in the well mixed case itself highlights the challenge in creating a true well mixed state in real spaces.

**Figure 31.**
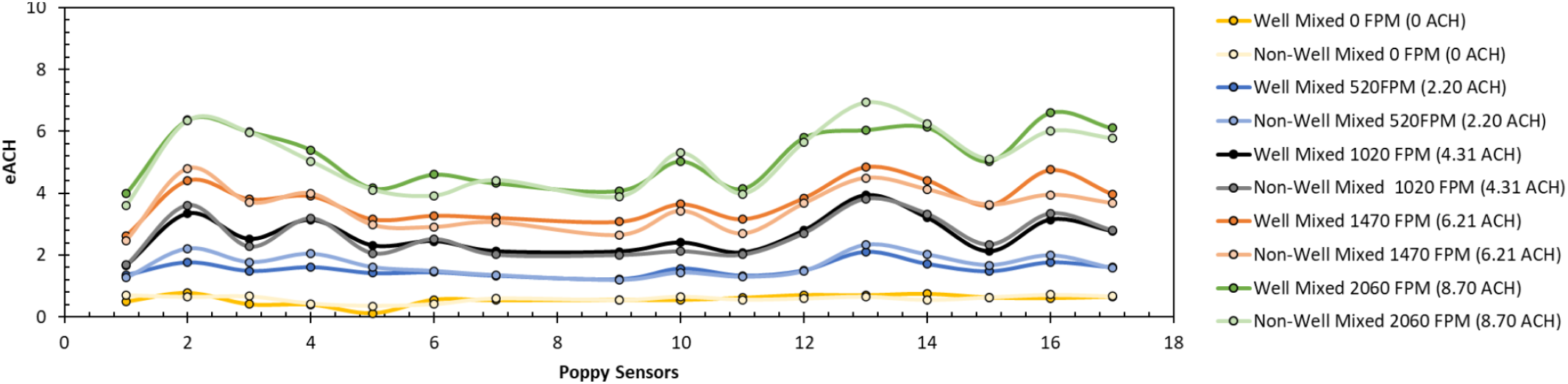
Positional spatial variation of eACH for a well mixed and a non-well mixed case for four different volumetric flow rates in the physical test space. Spatial variation is observed closer to the inlet, outlet, and walls.

Based on these findings, the placement of sensors in a space must consider the location of existing air vents, occupancy hotspots, walls, and how the space is used.

For each of the tests reported so far, the eACH calculation made use of the full decay period until the point that the concentration of tracer particles had returned to background levels. The time required for a tracer particle peak to decay to background depends on the air change rate, since higher air change rates remove more particles per unit time to reach initial conditions. The length of the tests conducted so far in this study using a “decay to background” method ranged from a span of several minutes for high eACHs, to hours for very low eACHs.

There are practical reasons why it is attractive to be able to shorten test time to a standard duration that can be conducted quickly and with confidence in typical indoor spaces. We reanalyzed the data from the cases reported above and found that a 15 min sampling time allows near-field nebulization effects to dissipate. A 15 min “rapid test” eACH corresponds closely to eACHs calculated using a complete decay period, while decreasing the decay sampling period to times shorter than 15 minutes leads to increased noise and a significant loss of accuracy. An average deviation of 4.97 % was observed for an eACH calculated using a 15 min sampling decay compared to the same using a full decay period. Figure 32 shows the relationship for eACH measurements using a full decay period compared to a 15-min rapid test, under the two levels of mixing at increasing volumetric flow rates.

**Figure 32.**
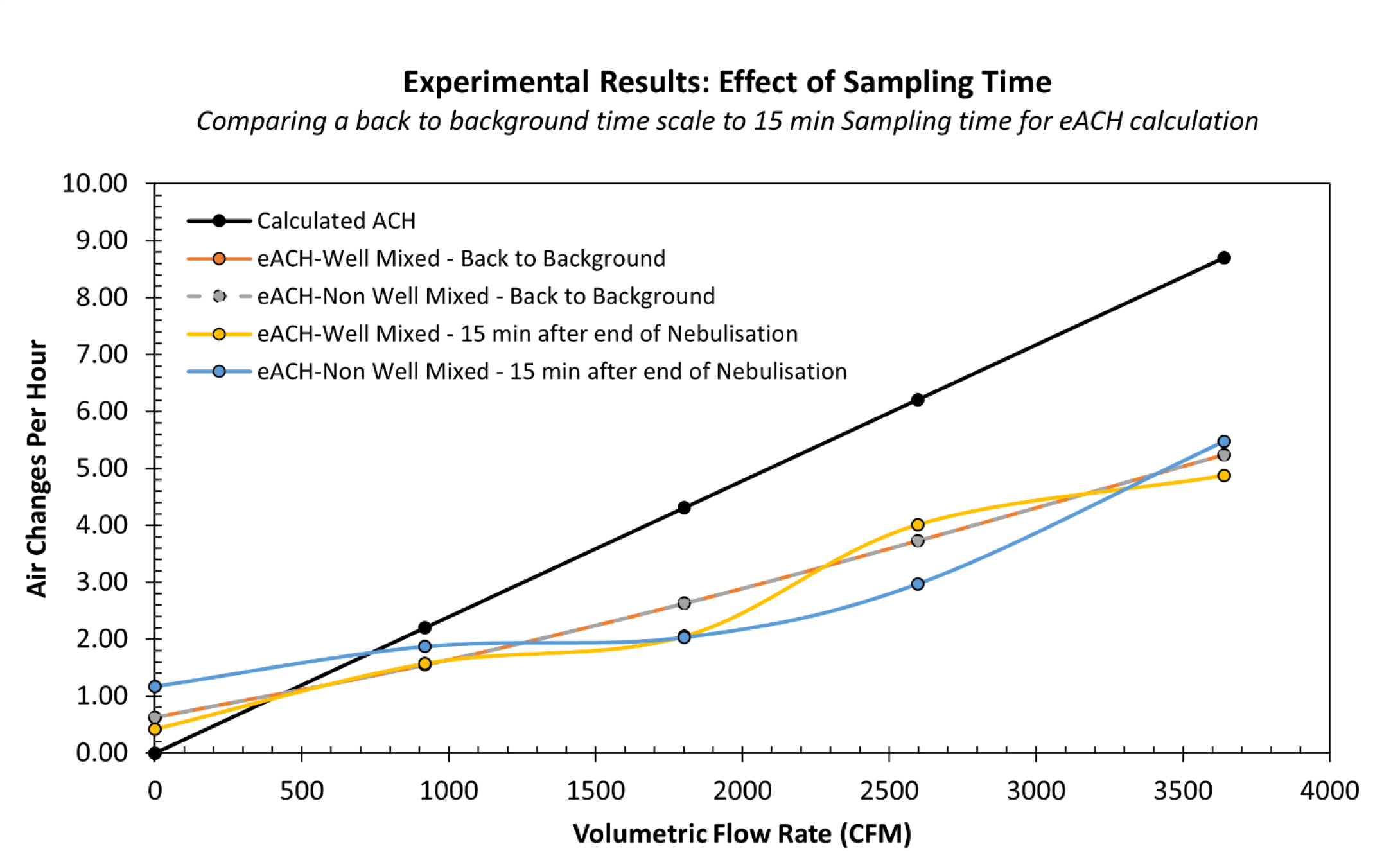
Effect of decay sampling time on the computation of eACH for all physical test cases.

## 6. Conclusions

In this study, we investigated the relationship between the calculated air change rate of a space (i.e. volumetric airflow based) and the effective air change rate for aerosol particle removal within the breathing zone based on direct measurements of tracer particle concentrations at representative occupant locations in a room. Further, we examined positional effects under well mixed and non-well mixed conditions, and determined a minimum decay sampling time needed for eACH measurements.

Using both CFD modeling and physical experiments we measured the decay rate of tracer aerosol particles in a large (25093 ft^3^) test space with an array of Poppy Sensors (virtual and physical) organized in a concentric zonal model around a nebulizer, with varying volumetric airflow rates. We focused on 1 μ*m* particles in the breathing zone (3 feet above the ground) as this size of aerosol represents a key peak in the distribution of respiratory aerosols produced by humans; aerosols of this size are known to remain airborne for long periods.

The two test beds for these analyses allowed us to compare results for eACH measurements under varying levels of control. The CFD model enabled a direct comparison of the differences between measured eACH and calculated ACH values under different levels of mixing, and provided airflow velocity visualization that could explain positional differences in eACH measurements.

Complementing this, physical experiments using nebulized DNA tracer particles and Poppy Sensors were used to validate the CFD results under real-world conditions that include all mechanisms of particle removal that contribute to true aerosol clearance rates, including deposition and leakage. In the physical study, a total of 5 relevant air change rate levels were tested under two conditions.

The key findings of this study are:

● The effective Air change Rate for fine respiratory aerosols in a non-well mixed room will differ (+/- 20-40%) from the ACH calculated from supply air. This is to be expected and the difference between the two metrics is fundamental to the system.

○ In rooms, this variance may be driven by static pressure effects, local heterogeneities and recirculation zones, and how airflow differs at the vent compared to the breathing zone where the tracer particles are nebulized (and where airborne transmissions occur). In addition to this, there are potentially accuracy issues in measuring volumetric flow rates using typical methods. Our results here are in agreement with the published literature (<u>19</u>).
● The magnitude and direction of the differences between eACH and ACH are challenging to anticipate based on prior knowledge of a room’s ventilation design and mixing level.
● Mixing, as performed in these tests, had little effect past a certain threshold – we observed similar eACH results for well mixed and non-well mixed cases in the physical tests.

○ Mixing was induced in the physical test space with the goal of producing a similar concentration of tracer particles across its volume. While localized mixing zones can lead to non-uniformities in a more segmented space (cubicles, separating dividers, shelves, etc.), additional mixing in the test space had little impact on eACH variability.
● Positional eACH variation is dependent on local airflow velocity. This has important implications for occupant safety, and use of zones within rooms. It also defines the need to select eACH measurement positions that are both representative of typical occupant usage and that are not heavily influenced by nearby vents, air purifiers, or other structures that concentrate airflows. The latter can be usually accomplished through visual inspection of a facility.
● A shorter decay sampling time yields similar eACH results as a full decay sampling time (i.e. complete decay to background), with a small degree of deviation (4.97 % average deviation). This enables eACH measurements to be executed using a standardized “rapid test” procedure that can be accomplished with a 15 minute decay sampling time.

Ventilation is one of the most critical components in a layered approach toward reducing the spread of airborne infectious diseases in indoor spaces. However, building ventilation systems act together with natural ventilation, local filtration systems and other aerosol removal processes to remove infectious aerosols from an occupied space. Therefore, measuring the particle removal rate in a space is a more direct measurement of the space’s relative safety from aerosol hazards than measurements of HVAC ventilation rates or CO2 measurements. The effective Air Change Rate (eACH) is a metric that estimates the building ACH using measurements of particle decay in a space. While it is only an estimate of ACH, it is a direct measurement of particle removal in a space, which, if measurements are made in the breathing zone, are also more directly related to the exposure of individuals in the space to infectious aerosols than measurement of ACH.

Here we have demonstrated that the eACH for a functional zone within space can be measured using a procedure that can be executed in any building and takes into account all active and passive aerosol particle removal mechanisms. The process involves the point release of liquid tracer particles containing DNA, and monitoring the decay in their local concentration using sensors that can accurately detect and count tracer particles within the 1μ*m* size fraction. This process is amenable to full automation, using wifi connected nebulizers and sensors that can be installed within a room, or executed on the fly using a portable “rapid test” kit. Further, these tests have demonstrated that simple duct face velocity measurements, used to estimate ACH, may overestimate the expected particle removal rate in the breathing zone of the space. The simple, automatable, process described here, provides a direct and more accurate estimation of occupant safety in a space, which is inclusive of all particle removal processes.

The availability of a direct test for eACH enables empirical optimization of the combination of ventilation and filtration mechanisms in any room to reach and maintain target aerosol clearance rates that deliver reliable airborne infection control. Importantly, tests of the effective air change rate (eACH) are direct measurement of the equivalent air changes per hour (ACHe) added from outdoor air. Given ASHRAE’s 2023 commitment to develop a new indoor air quality standard for pathogen mitigation (Standard 241), and the existence of an estimated >1T square feet of commercial indoor space in the world, new rapid, accurate and scalable technologies for ventilation verification such as this will be needed for any ACHe-based standard to be efficiently adopted by buildings on a national or global scale.

## Data Availability

All data produced in the present study are available upon reasonable request to the authors.

## Appendix A

### Modeling of Particle Size Distribution in ANSYS Fluent

#### Theory

The Rosin Rammler particle size distribution function is one of the well-known distribution functions and is applicable in various settings like development of spray technologies, aerosol science, emulsification processes etc. It is also used to investigate the effects of the particle size distribution on the ignition energy of explosive dusts [3]. The cumulative form for the Rosin – Rammler particle size distribution is defined by the following equation:

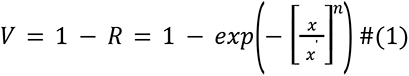

where,

x = particle size (x ≥ 0) expressed in, e.g., μ m

x′ = location parameter of the distribution, i.e., the particle size at a volume fraction of 0.368 oversize (x′ > 0)

V = mass or volume fraction of particles with sizes smaller than or equal to x (undersize distribution), assuming constant mass density of all particles

R = mass or volume fraction of particles with sizes larger than x (oversize distribution) n = spread parameter of the distribution (n > 0)

A Rosin-Rammler particle size distribution function was used to introduce particles of diameters ranging from 0.3 microns to 2 microns with a mean diameter of 0.7 microns. The particle size distribution plot for particles used in all the cases is shown in **Figure A1**.

**Figure A1.**
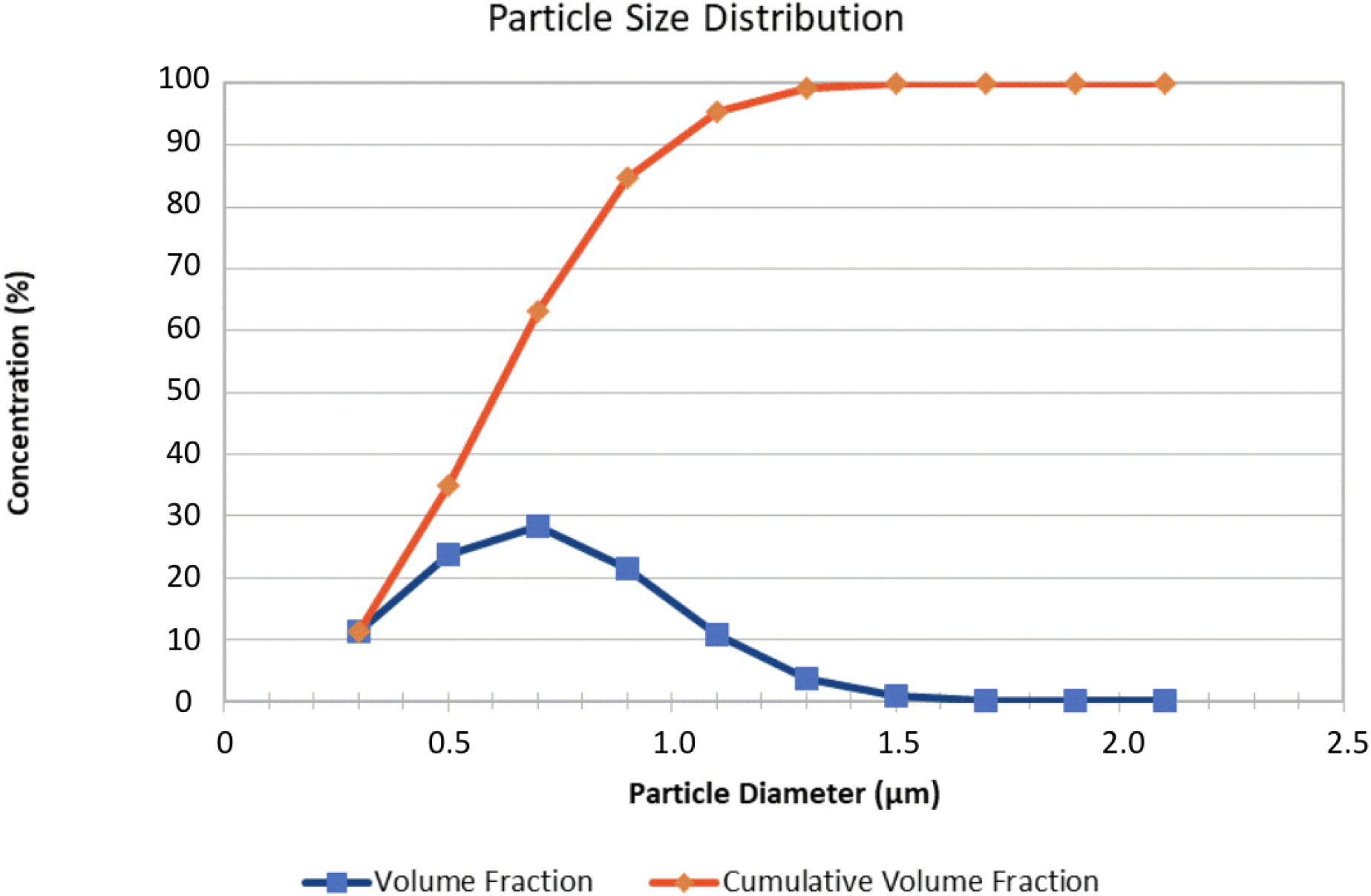
The particle size distribution used for all cases.

## Appendix B

All dimensions are in feet (ft).

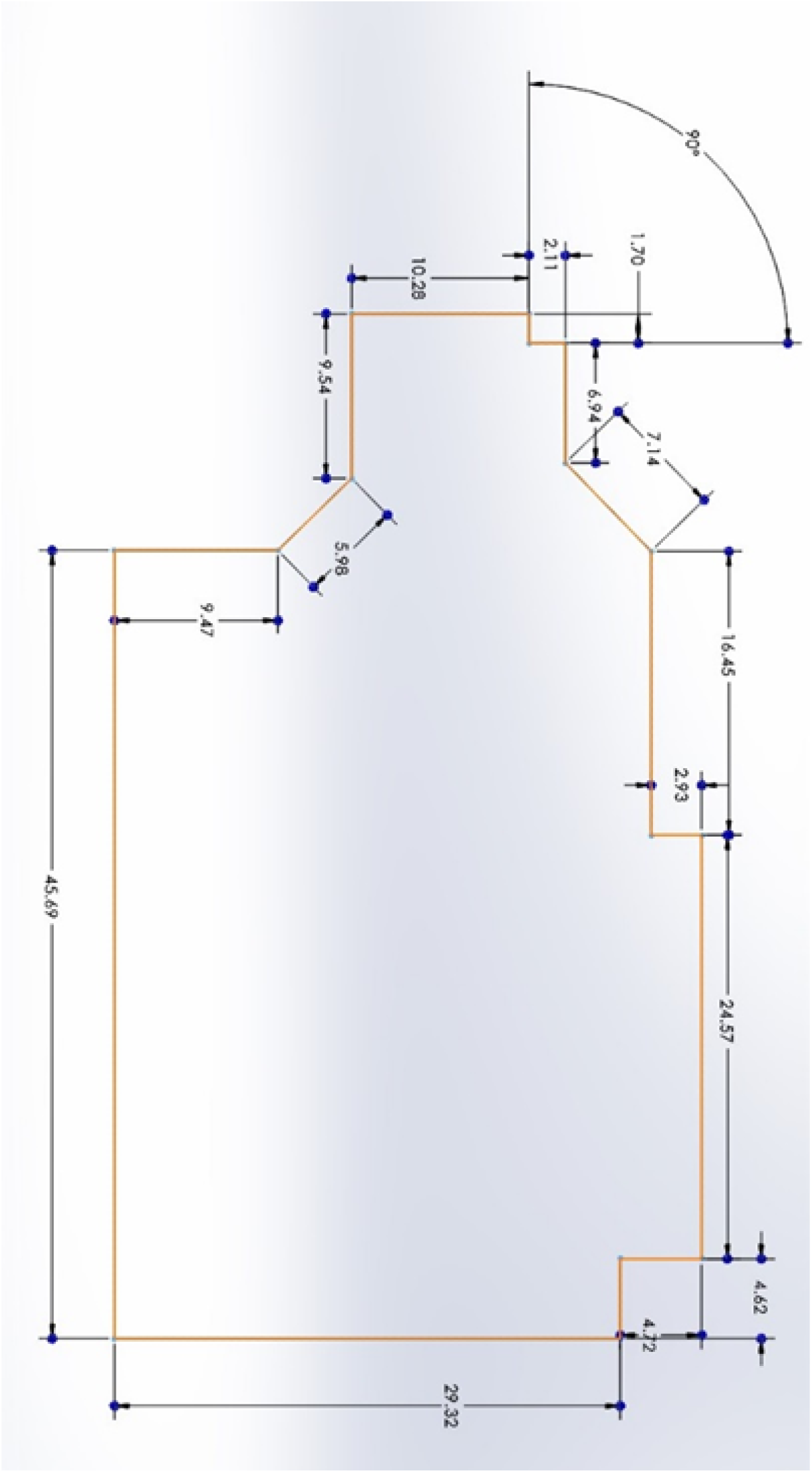

